# Characterizing the phenotypic and genetic structure of psychopathology in UK Biobank

**DOI:** 10.1101/2023.09.05.23295086

**Authors:** Camille M. Williams, Hugo Peyre, Tobias Wolfram, Younga H. Lee, Tian Ge, Jordan W. Smoller, Travis T. Mallard, Franck Ramus

## Abstract

Mental conditions exhibit a higher-order transdiagnostic factor structure which helps to explain the widespread comorbidity observed in psychopathology. However, the phenotypic and genetic structures of psychopathology may differ, raising questions about the validity and utility of these factors. Here, we study the phenotypic and genetic factor structures of ten psychiatric conditions using UK Biobank and public genomic data. Although the factor structure of psychopathology was generally genetically and phenotypically consistent, conditions related to externalizing (e.g., alcohol use disorder) and compulsivity (e.g., eating disorders) exhibited cross-level disparities in their relationships with other conditions, plausibly due to environmental influences. Domain-level factors, especially thought disorder and internalizing factors, were more informative than a general psychopathology factor in genome-wide association and polygenic index analyses. Collectively, our findings enhance the understanding of comorbidity and shared etiology, highlight the intricate interplay between genes and environment, and offer guidance for psychiatric research using polygenic indices.

## Main

Psychiatric disorders are among the leading contributors to the global disease burden. They are the main cause of years lived with disability and will affect more than 25% of the world’s population at some point in their lifetime. Critically, these disorders often co-occur with one another^1^, which further exacerbates the burden on individuals, their families, and communities^2,3^. Indeed, complex patterns of comorbidity are perhaps the norm in psychopathology, as more than 50% of people who meet diagnostic criteria for one disorder will also meet criteria for a second^1^.

In response to these observations of pervasive comorbidity, researchers in clinical psychology and psychiatry have increasingly turned toward transdiagnostic approaches in the study of mental conditions^4,5^. By cutting across fuzzy diagnostic boundaries, transdiagnostic research aims to identify and understand core mechanisms in psychopathology^6–8^. Such insights are uniquely poised to reduce the burden of mental illness, as they may advance knowledge of multiple disorders simultaneously, contribute to the development of therapeutics with broad utility, and even inform future nosological systems in the field^9–11^.

Advances in statistical and psychiatric genetics have substantiated this fundamental change in perspective, especially for biological investigations. Results from large-scale genome-wide association studies (GWAS) have confirmed that all forms of psychopathology are partly heritable^11,12^ and highly polygenic – influenced by thousands of causal variants with probabilistic effects^9,13,14^. Notably, many disorder-linked variants appear to have pleiotropic effects, conferring risk in a transdiagnostic manner^11^. With overlap at the genetic level typically mirroring that at the phenotypic level^15^, these patterns of widespread pleiotropy support the hypothesis that comorbidity among disorders may, in part, arise from shared etiology.

Concurrently, factor analytic research has revealed a transdiagnostic structure of psychopathology that is observed at both phenotypic and genetic levels^11,16–21^. Generally, these studies report that individual disorders comprise higher-order factors of internalizing, externalizing, and thought disorder problems that help explain comorbidity^17,22^. They even suggest that a general transdiagnostic dimension – the ‘*p*’ factor – explains features that are common to all mental disorders^16,23,24^. However, recent data-driven genomic studies report genetic factor structures that differ from the phenotypic literature, raising questions about the utility and interpretation of the *p* factor^9^. In fact, one recent genomic study found that the *p* factor produced few biological insights, as it obfuscated patterns of associations with individual variants, genetic correlations with biobehavioral outcomes, and enrichment within specific biological annotations^9^.

Here, we aim to advance the understanding of psychiatric comorbidity with two extensive sets of analyses. First, we report a comprehensive factor analysis of 10 psychiatric conditions in the UK Biobank, and we characterize model (dis)similarity across phenotypic and genetic levels of analysis. Second, we evaluate the utility of the *p* factor in biological investigations of comorbidity, using GWAS and polygenic indices (PGIs) as exemplars. In doing so, our results provide substantive insights into the factor structure of psychopathology and practical guidance on how to leverage these insights.

## Results

In the present study, we investigated the factor structure of 10 diverse psychiatric conditions in the UK Biobank: generalized anxiety (ANX), alcohol use disorder (AUD), bipolar disorder (BIP), depression (DEP), eating disorder (EAT), obsessive-compulsive disorder (OCD), posttraumatic stress disorder (PTSD), schizophrenia (SCZ), suicidality (SUI), and substance use disorder (SUD). As described in the **Methods**, these dichotomous outcomes were constructed using self-report and diagnostic measures (**Supplementary Tables 1-2**). To facilitate GWAS and PGI analyses, we divided the UK Biobank into discovery and holdout samples (see **Methods**). Analyses were conducted at phenotypic and genetic levels, which are denoted with _p_ and _g_, respectively (e.g., *r*_p_ = phenotypic correlation, *r*_g_ = genetic correlation).

To investigate the phenotypic *and* genetic factor structure of psychiatric conditions, we first conducted univariate genome-wide association analyses (GWASs) for the 10 psychiatric conditions in up to 402,411 UK Biobank participants. However, as some phenotypes had relatively few cases in UK Biobank (**Fig. 1a**), we performed genomic meta-analyses using publicly available data to improve statistical power for downstream analyses (**Methods**). UK Biobank phenotypes were sufficiently similar to those from external cohorts (**Supplemental Information**) and cross-cohort genetic correlations were generally moderate to high (**Supplementary Table 3**). The difference in the genetic correlations of psychiatric conditions from the UK Biobank and the meta-analyzed GWASs tended to be small to moderate (median |Δ*r*_g_| = 0.073); **Supplementary Fig. 1**). Therefore, we report genetic results from the meta-analyzed GWASs in the main text.

**Figure 1.**
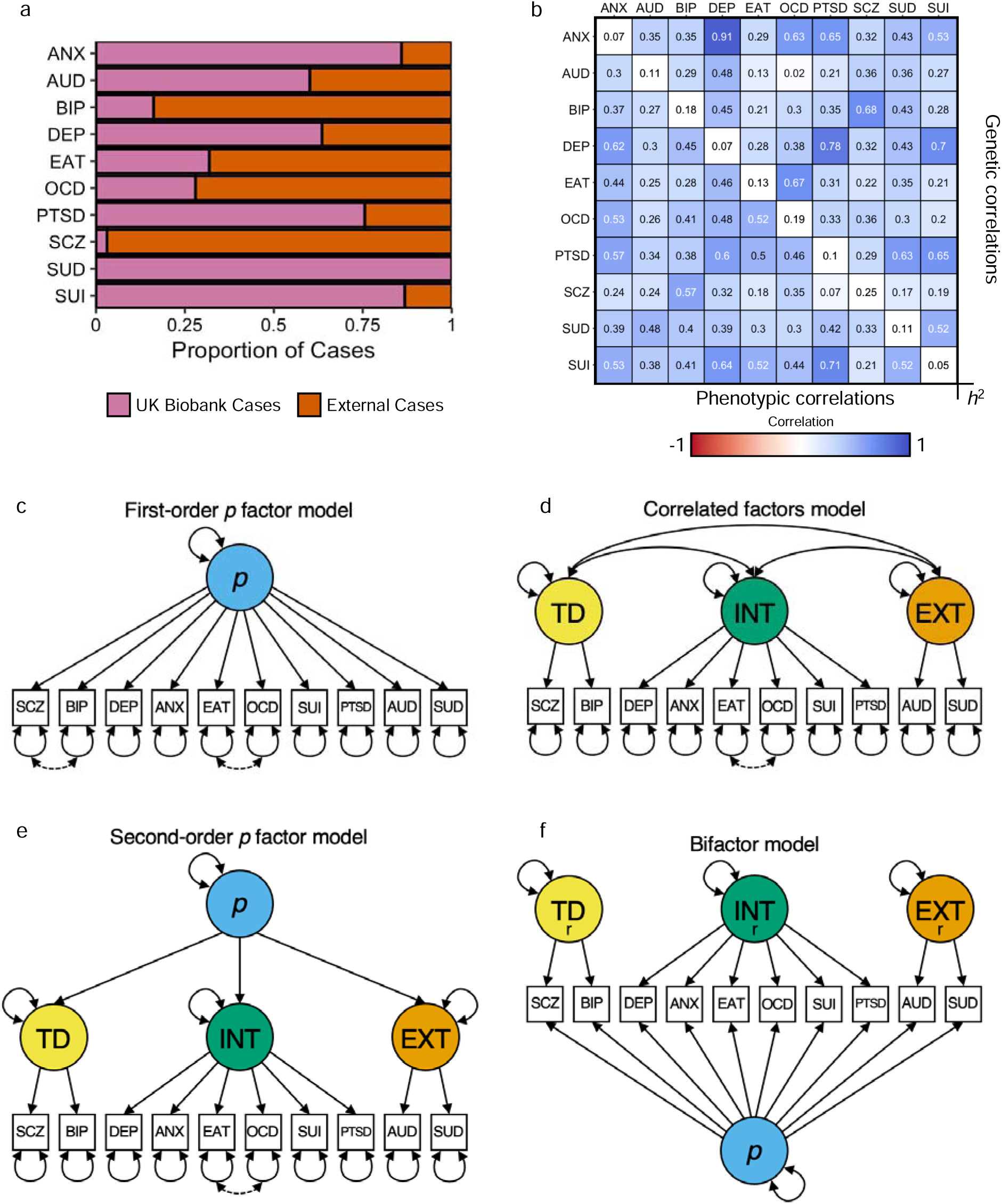
Overview of the data and models tested in the present study. **a**, Bar chart illustrating the proportion of cases in the GWAS meta-analyses from the UK Biobank versus external cohorts. **b**, A matrix of the phenotypic and genetic correlations among psychiatric conditions. Heritability on the observed scale is reported along the diagonal, while phenotypic and genetic correlations are reported in the lower and upper triangles, respectively. Genetic correlations were estimated with LD score regression^25^. **c-f**, Path diagrams of the factor models tested in the present study. Solid lines reflect associations across phenotypic and genetic models, while dotted lines reflect residual correlations that are specific to the genetic models. ANX = Generalized anxiety, AUD = alcohol use disorder, BIP = bipolar disorder, DEP = depression, EAT = eating disorder, OCD = obsessive-compulsive disorder, PTSD = posttraumatic stress disorder, SCZ = schizophrenia, SUI = suicidality, and SUD = substance use disorder. r = residualized factor after taking into account variance shared across all disorders (*p* factor).

All psychiatric conditions were significantly heritable (*h*^2^ = 0.057 – 0.476; **Table 1**) and test statistics showed substantial inflation (λ_GC_= 1.023 – 1.699, mean χ^2^ = 1.03 – 1.955; **Table 1**), indicative of a robust polygenic signal for all phenotypes. Linkage disequilibrium (LD) score regression intercepts and attenuation ratios indicated that these signals were primarily attributable to polygenic architectures rather than population stratification or other confounding (intercept = 1 – 1.083, attenuation ratio = less than 0 – 0.157; **Table 1**).

**Table 1.** Summary of the 10 psychiatric conditions included in the present study.

| Phenotype<br>(abbreviation) | Phenotypic Analyses |  |  |  | Genetic Analyses |  |  |  |  |  |  |  |  |
| --- | --- | --- | --- | --- | --- | --- | --- | --- | --- | --- | --- | --- | --- |
|  | N | Sample Lifetime Prevalence |  |  | UK Biobank<br>GWAS | Public GWAS |  | Meta-analyzed GWAS |  |  |  |  |  |
| | | Full | CFA | Prediction | $N_{eff}$ | Sum of<br>$N_{eff}$ | Ref. | $Max$<br>$N_{eff}$ | $h^2$ (SE) | $\lambda_{GC}$ | Mean $\chi^2$ | Intercept<br>(SE) | Ratio (SE) |
| Alcohol Use Disorder (AUD) | 465,858 | 3.16% | 3.22% | 3.96% | 49,557 | 23,282 | 26 | 72,839 | 0.114<br>(0.011) | 1.124 | 1.147 | 1.023<br>(0.008) | 0.157<br>(0.055) |
| Generalized Anxiety (ANX) | 465,086 | 10.01% | 11.14% | 16.16% | 147,311 | 14,806 | 27 | 168,782 | 0.071<br>(0.005) | 1.210 | 1.229 | 1.020<br>(0.009) | 0.087<br>(0.038) |
|  |  |  |  |  |  | 6,665 | 28 |  |  |  |  |  |  |
| Bipolar Disorder (BIP) | 465,148 | 1.04% | 1.04% | 1.09% | 15,629 | 46,582 | 29 | 62,211 | 0.185<br>(0.009) | 1.320 | 1.385 | 1.022<br>(0.008) | 0.056<br>(0.020) |
| Depression (DEP)** | 465,211 | 19.57% | 24.32% | 28.98% | 254,389 | 111,221 | 30 | 365,610 | 0.069<br>(0.003) | 1.3409 | 1.412 | 1.015<br>(0.001) | 0.037<br>(0.024) |
| Substance Use Disorder (SUD)* | 465,007 | 0.79% | 0.80% | 1.23% | 11,934 | - | - | 11,454 | 0.109<br>(0.051) | 1.023 | 1.030 | 1.003<br>(0.008) | 0.091<br>(0.260) |
| Eating Disorders (EAT) | 465,010 | 0.42% | 0.42% | 0.78% | 6,513 | 10,164 | 31 | 16,677 | 0.131<br>(0.019) | 1.071 | 1.079 | 1.000<br>(0.008) | Ratio < 0 |
| Obsessive-Compulsive Disorder (OCD) | 465,052 | 0.26% | 0.26% | 0.39% | 4,173 | 7,281 | 32 | 11,454 | 0.187<br>(0.033) | 1.059 | 1.059 | 1.000<br>(0.007) | 0.001<br>(0.120) |
| Post-Traumatic Stress Disorder (PTSD) | 465,052 | 1.93% | 1.94% | 3.98% | 29,464 | 5,831 | 33 | 35,295 | 0.097<br>(0.017) | 1.065 | 1.070 | 1.004<br>(0.008) | 0.058<br>(0.110) |
| Schizophrenia (SCZ) | 465,058 | 0.37% | 0.38% | 0.10% | 5,183 | 99,863 | 34 | 105,033 | 0.246<br>(0.011) | 1.699 | 1.955 | 1.083<br>(0.012) | 0.087<br>(0.012) |
| Suicidality (SUI) | 465,069 | 9.86% | 11.01% | 21.64% | 143,212 | 21,208 | 35 | 164,420 | 0.048<br>(0.003) | 1.197 | 1.219 | 1.013<br>(0.008) | 0.060<br>(0.038) |

### Factor analyses reveal the dimensional structure of psychopathology in UK Biobank

To better understand patterns of comorbidity and shared etiology among psychiatric conditions, we calculated the pairwise phenotypic and genetic correlations (*r*_p_ and *r*_g_) for all study phenotypes (**Fig. 1b**). In both matrices, we observed a positive manifold among mental conditions, with modest-to-large positive correlations across most pairwise combinations (mean *r*_p_ = 0.404 [range = 0.074 – 0.710], mean *r*_g_ = 0.39 [range = 0.022 – 0.908]). We then used exploratory and confirmatory factor analyses (EFA and CFA, respectively) to characterize the multivariate system of relationships between psychiatric conditions at phenotypic and genetic levels.

Guided by the results of a parallel analysis (**Supplementary Fig. 4a**), we modeled up to five phenotypic latent factors using EFA. The phenotypic covariance among conditions was parsimoniously described with three factors, as the Δ*R*^2^ and change in root mean square error of approximation (ΔRMSEA) between the three- and four-factor models were less than 0.05 and 0.015, respectively. We observed a similar pattern in the genetic data, where the inclusion of a fourth factor did not substantially increase the variance explained over the three-factor model (Δ*R*^2^ = 0.06; **Supplementary Fig. 4b**). Notably, the phenotypic and genetic models both exhibited approximate simple structure with negligible cross-loadings for most psychiatric conditions (**Supplementary Table 5**). However, cross-level differences in factor loadings suggested that the structure of psychopathology may partially differ across phenotypic and genetic models.

In the phenotypic EFA, one factor was defined by thought disorder conditions (BIP and SCZ), another factor by internalizing conditions (ANX, DEP, EAT, OCD, PTSD, and SUI), and a final factor by externalizing/substance use conditions (AUD and SUD). In the genetic EFA, we observed a similar thought disorder factor (BIP and SCZ), but the internalizing factor now included substance use conditions (ANX, AUD, DEP, PTSD, SUD, and SUI) and the third factor was now composed of conditions characterized by compulsivity (EAT and OCD). The full results are reported in **Supplementary Table 5**.

Informed by psychometric theory and the above results, we fit four CFA models to the phenotypic and genetic data: (i) a first-order factor model, (ii) a correlated factors model with three latent factors; (iii) a second-order factor model, where a second-order *p* factor of psychopathology explains variance in first-order psychopathology factors; and (iv) a bifactor model, where four orthogonal factors (one general *p* factor, three domain-specific factors) explain variance in psychiatric conditions (see illustrative path diagrams in **Fig. 1c-f**; see **Supplementary Fig. 5-6** and **Supplementary Tables 6-8** for path diagrams with parameter estimates).

While all models showed relatively good fit within their respective level of analysis (all models: comparative fit index [CFI] > 0.971 [range = 0.961 – 0.980] and standardized root mean square residual [SRMR] < 0.083 [range = 0.079 – 0.086]), the correlated factors and second-order factor models optimized the balance between parsimony and goodness-of-fit^37^ in the phenotypic and genetic data.

### Cross-level disparities reveal gene-environment interplay in comorbidity & shared etiology

As our EFAs suggested differences in the phenotypic and genetic factor structure of psychopathology, we sought to more formally characterize these dissimilarities. Specifically, we aimed to identify which psychiatric conditions contribute to this discordance and probe the extent to which measurement models were specific to phenotypic and genetic levels of analysis.

Although the phenotypic and genetic correlation matrices were highly correlated (*r* = 0.873, *p* = 9.99e-4, several pairwise relationships differed across levels of analysis. To quantify these dissimilarities, we calculated the ‘disparity’^38,39^ between bivariate phenotypic and genetic correlations (*D* = *r*_p_ – *r*_g_) (**Fig. 2a**). Disparities were generally modest overall (mean |*D*| = 0.104), though there were considerable deviations (range *D* = -0.290 – 0.317) (**Fig. 2b**). For example, SUI had larger phenotypic than genetic correlations (mean *D*_SUI_ = 0.091), and exhibited large disparities with compulsive conditions (*D*_SUI,EAT_ = 0.317; *D*_SUI,OCD_ = 0.247). Conversely, DEP had smaller phenotypic than genetic correlations (mean *D*_DEP_ = -0.051), and showed some marked differences with frequently comorbid conditions (*D*_DEP,ANX_ = -0.29; *D*_DEP,PTSD_ = -0.182; *D*_DEP,AUD_ = -0.181). Notably, observed disparities were not systematically related to imprecision (standard errors) in genetic correlation estimates (*r* = -0.001, *p* = 0.992, **Fig. 2c**), the proportion of external cases in the GWAS data (all *p* > 0.05), or the GWAS Neff (only ANX and AUD *p* < 0.05, all other *p* > 0.05) (**Methods**). This suggests that differences might reflect environmental influences causing phenotypic correlations to deviate from genetic ones.

**Figure 2.**
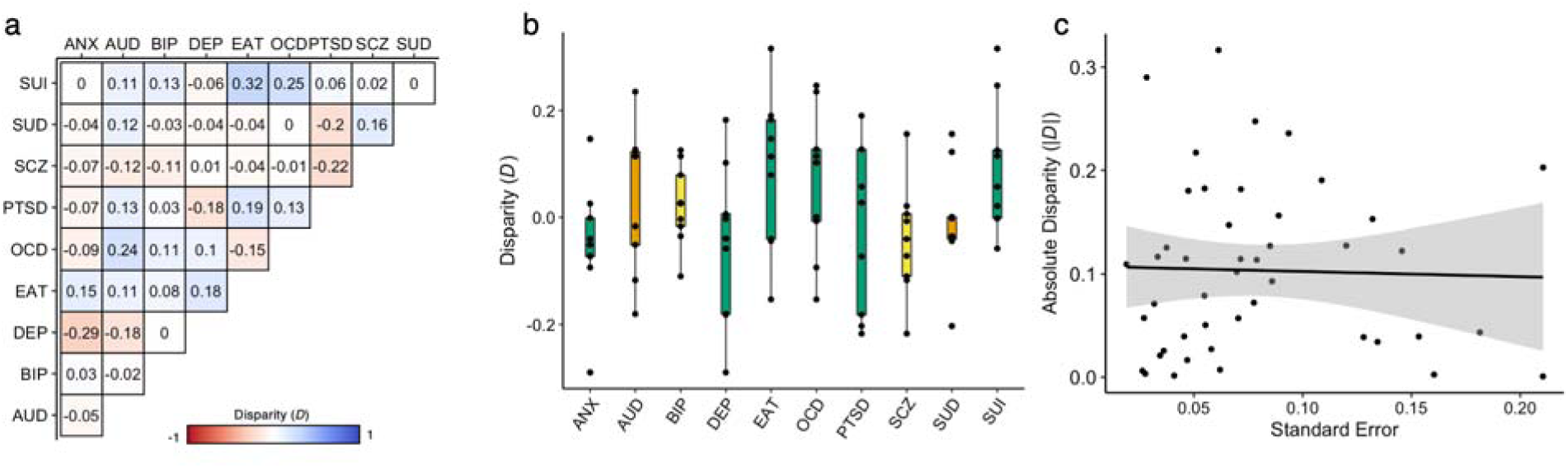
Comparison of phenotypic and genetic correlations among 10 psychiatri conditions. **a**, Disparity between the phenotypic and genetic correlations for each pairwise combination of psychiatric conditions (*D* = *r*_p_ -*r*_g_). **b**, Box plots illustrating the distribution of *D*, summarized for each condition. Box plots are colored according to the corresponding factor from the phenotypic model. **c**, Scatter plot of the association between |*D|* and the imprecision of the genetic correlation estimate, as indexed by the standard errors. ANX = generalized anxiety, AUD = alcohol use disorder, BIP = bipolar disorder, DEP = depression, EAT = eating disorder, OCD = obsessive-compulsive disorder, PTSD = posttraumatic stress disorder, SCZ = schizophrenia, SUI = suicidality, and SUD = substance use disorder.

To evaluate the degree to which the structure of psychopathology may be level-specific, we fit the best phenotypic and genetic models from the EFAs to the phenotypic and genetic data (**Fig. 3a-d**; **Supplementary Table 9**). We found that the phenotypic model showed acceptable fit to the genetic data (CFI = 0.969, SRMR = 0.106), which further improved when the EAT and OCD residuals were allowed to correlate (CFI = 0.975, SRMR = 0.083; **Fig. 3d**). Conversely, the genetic model fit the phenotypic data well with no need for additional parameters (CFI = 0.992; SRMR = 0.055; **Fig. 3c**). However, the correlation between the compulsive and internalizing factors was 0.889, suggesting that these factors are much more similar phenotypically than genetically (*D*_INT_,_COMP_ = 0.449; **Fig 3b**). Note that the loadings of factors with the same indicators across phenotypic and genetic models were highly consistent (*r* = 0.998 and 0.999 for the loadings across models for the phenotypic and genetic data, respectively). Sensitivity analyses suggest these results were robust to several potential confounds, such as sampling differences across datasets (**Supplemental Information**).

**Figure 3.**
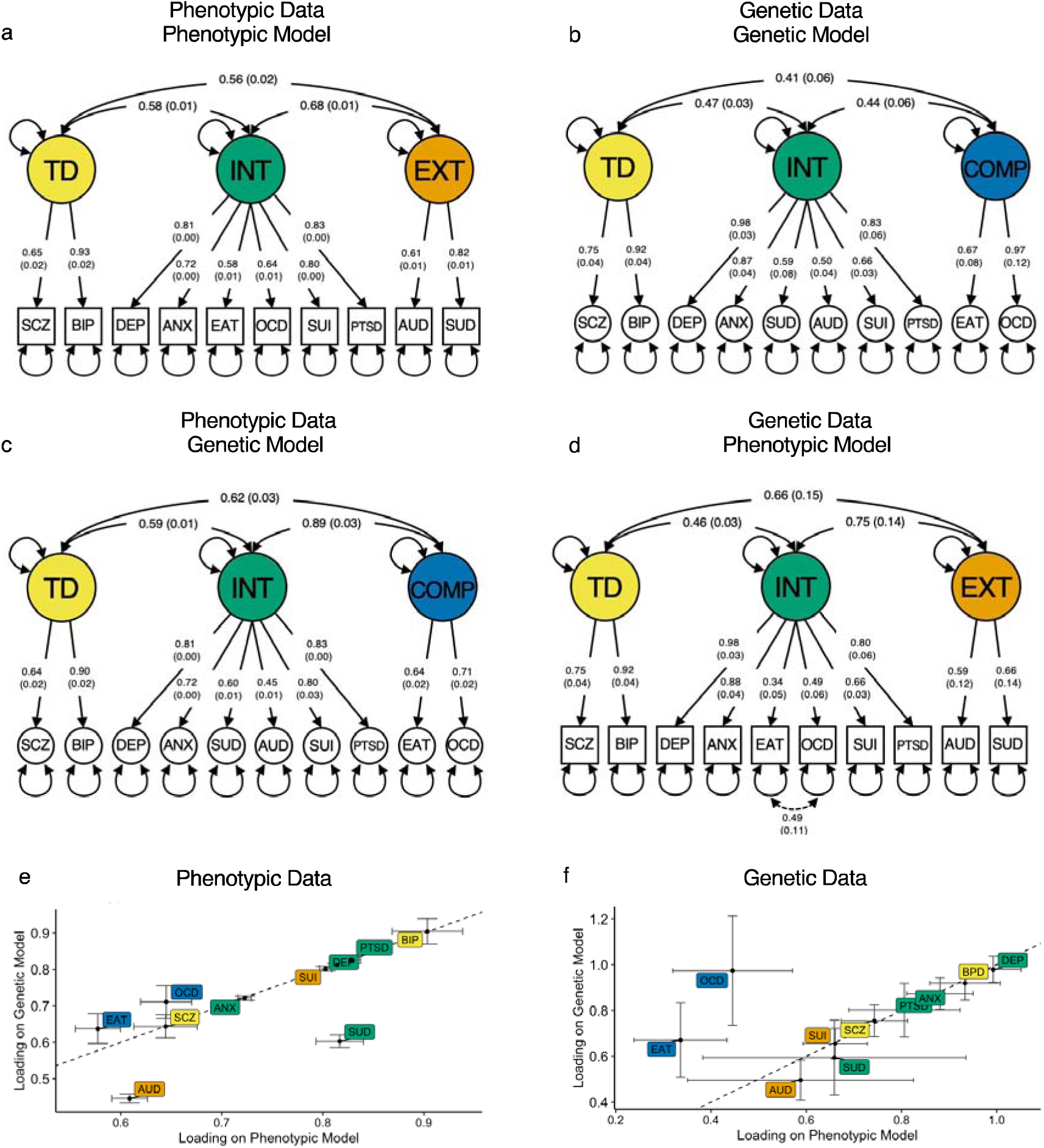
Cross-level comparison of the phenotypic and genetic factor structure of psychopathology. **a-d**, Path diagrams of the best-fitting phenotypic and genetic correlated factors model on the phenotypic data from the UK Biobank and genetic data from the meta-analyses of genome-wide analysis studies. Residual variances were omitted for plotting purposes (see **Supplementary Tables S6 and S8** for further information, respectively). **e**, Scatter plot of the loadings of the phenotypic (**a**) and genetic (**c**) correlated factors model applied to the phenotypic data. **f**, Scatter plot of the loadings of the phenotypic (**d**) and genetic (**b**) correlated factors model applied to the genetic data. The dotted line reflects a perfect correlation (slope of 1). TD = thought disorder factor, INT = internalizing factor, COMP = compulsive disorders factor, EXT = externalizing factor, ANX = generalized anxiety, AUD = alcohol use disorder, BIP = bipolar disorder, DEP = depression, EAT = eating disorder, OCD = obsessive-compulsive disorder, PTSD = posttraumatic stress disorder, SCZ = schizophrenia, SUI = suicidality, and SUD = substance use disorder.

Collectively, these results suggest that the factor structure of psychopathology is quite similar across levels of analysis for thought disorder and internalizing but may differ for compulsive and externalizing disorders, possibly due to environmental influences.

### Domain-level transdiagnostic factors show robust validity and utility in genetic analyses

To investigate the validity of the transdiagnostic factors, we conducted GWAS and *Q*_SNP_ analyses. The *Q*_SNP_ analysis is particularly useful in this regard, as it indexes the degree of heterogeneity observed in the SNP-level effects by testing whether SNPs plausibly operate through the factor as opposed to directly on the indicators (**Methods**). We performed these analyses for eight latent factors in Genomic SEM^36^: the correlated factors from the phenotypic model (TD_p_, INT_p_, and EXT_p_), the correlated factors from the genetic model (TD_g_, INT_g_, and COMP_g_), and two versions of a general psychopathology factor from the phenotypic models: one from a first-order factor model and one from a second-order factor model (Fig. 1c**,e**; first-order *p* and second-order *p*; **Supplemental Information** for further details on the bifactor model). We did not run GWAS for both genetically and phenotypic versions of the second-order *p* factor, as results demonstrated that slight configural changes did not affect GWAS results for other first-order factors (see **Supplemental Information**). For example, internalizing and thought disorder factors from the phenotypic model were highly correlated (*r*_g_ > 0.992) with their respective factors in the genetic models, as were *p* factors across models (*r*_g_ = 0.985; **Supplementary Fig. 14**).

We observed substantial inflation of the GWAS test statistics for all latent genetic factors (λ_GC_= 1.091 – 1.607, mean GWAS χ^2^ = 1.056 – 1.809), which was primarily attributable to polygenic architecture (intercept = 0.955 – 1.001, attenuation ratio = less than 0 – 0.001; **Table 2**). With respect to the *Q*_SNP_ statistics, we found relatively low degrees of heterogeneity for the domain-specific transdiagnostic factors (TD_p_, INT_p_, EXT_p_, TD_g_, INT_g_, and COMP_g_), which indicates that many of the associated loci plausibly operate through a common pathway (**Supplemental Information**). However, this was not true for the first- and second-order *p* factors, where we observed substantially greater inflation in the *Q*_SNP_ statistics (ratios of mean GWAS χ^2^ / *Q*_SNP_ χ^2^ = 0.987 and 1.020, respectively). Coupled with the low number of associated loci with either configuration of the *p* factor (**Table 2**, Fig. 3a**,b**), these results suggest that the common variants underlying the genetic architecture of psychopathology are not adequately captured by the *p* factor. Although locus discovery was not the aim of the present study, we report the significant genomic loci of each psychopathology factor in the supplement (**Supplementary Table 4**, **Methods**).

**Table 2.**
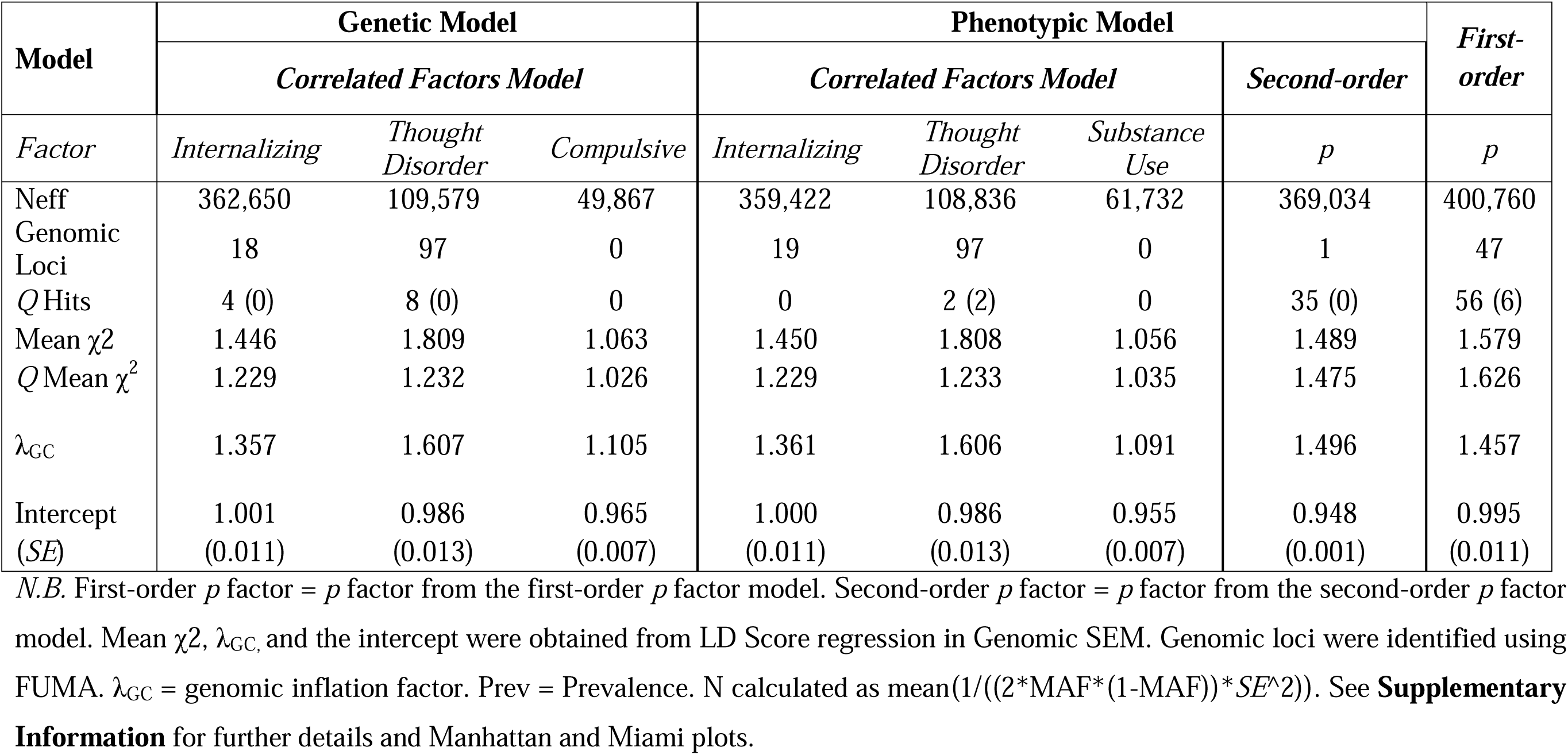
Results from multivariate genome-wide association analyses.

Comparison to models in the literature provided strong evidence of concurrent validity for our latent factors of psychopathology. For instance, our EXT factor was genetically correlated at 1 (*SE* = 0.148) with a well-powered externalizing factor model^40,41^. Similarly, our INT_p_ and TD_p_ factors were strongly correlated with genomic factors of internalizing and thought disorder psychopathology^42^, respectively (INT *r*_g_ = 0.903, *SE* = 0.028; TD *r*_g_ = 1, *SE* = 0.032). Although models from the literature included UK Biobank data, they also used data from other cohorts and included conditions that were not well represented in UK Biobank (e.g., attention-deficit/hyperactivity disorder for externalizing, schizoaffective disorder for thought disorder), supporting the generalizability of our inferences.

We further interrogated the validity of these factors via *Q_Trait_*analyses, which index heterogeneous effects at the level of genetic correlations by testing whether genetic relationships between psychiatric conditions and other complex traits plausibly operate through latent genetic factors. Once again, domain-level transdiagnostic factors showed low rates of heterogeneity while both configurations of the *p* factor exhibited substantial heterogeneity (**Supplemental Information**). For example, of the 38 phenotypes that were significantly genetically correlated with the second-order *p* factor, 31 (82%) were significant in the *Q_Trait_*analyses (**Supplementary Fig. 11**). In general, we found that genetic correlations between external traits and the *p* factor were similar to those with INT, but differed from those with TD (Fig. 3c), suggesting that the *p* factor may obfuscate domain-specific associations.

### Polygenic indices for the *p* factor obfuscate domain-specific associations

Finally, we examined whether the *p* factor provided additional utility over the domain-level factors in the context of PGI analyses. Here, we compared associations between transdiagnostic PGIs and outcomes related to psychopathology (8 factors) and neuroanatomy (62 cortical mean thickness and 62 surface areas) in a UK Biobank holdout sample of approximately 24,100 individuals (**Method**). We calculated PGIs for the eight factors described above: TD_p_, INT_p_, EXT_p_, TD_g_, INT_g_, COMP_g_, first-order *p*, and second-order *p*.

All PGIs were significantly associated with their cognate outcome (e.g., PGIs for TD_p_ and TD_g_ were associated with a phenotypic thought disorder factor) and most showed cross-trait patterns of association (**Supplemental Table 11**). With respect to the *p* factor PGIs, the second-order *p* factor PGI explained less variance than the combined three PGIs from the correlated factors model (e.g., TD_p_, INT_p_, and EXT_p_ or TD_g_, INT_g_, and CMP_g_) across all scenarios. This was also true for the first-order *p* factor PGI, except for compulsive disorders, where it explained more variance in compulsive disorders than the three PGIs from the correlated factors models (Δ*R*^2^ = 0.63 and 0.81%, respectively). Generally, the PGIs of the genetic and phenotypic correlated factors model of well-powered GWAS (i.e., TD and INT) tended to explained more or similar variance to the *p* factor PGIs (**Supplemental Table 11**).

When predicting individual differences in cortical thickness and surface area (Fig. 4d), we found that associations between PGIs and regional structural metrics were quite variable (**Supplementary Table 13**, **Supplementary Fig. 23-24**). For instance, the INT_p_ PGI was generally positively associated with cortical mean thicknesses, whereas the TD_p_ PGI tended to be negatively associated with cortical mean thicknesses (Fig. 4e). Consequently, these associations were often obfuscated when using a *p* factor PGI. Further examination revealed that the *p* factor results largely reflected diminished effects of the INT_g_ PGI – a pattern that was consistent for both first-and second-order *p* factor PGIs (correlations between INT_g_ and *p* factor effects ∼ 0.749; Fig. 4f). While we did not observe similarly strong patterns with surface area outcomes (**Supplementary Table 13**), this is likely a consequence of the very small effect sizes estimated in those models.

**Figure 4.**
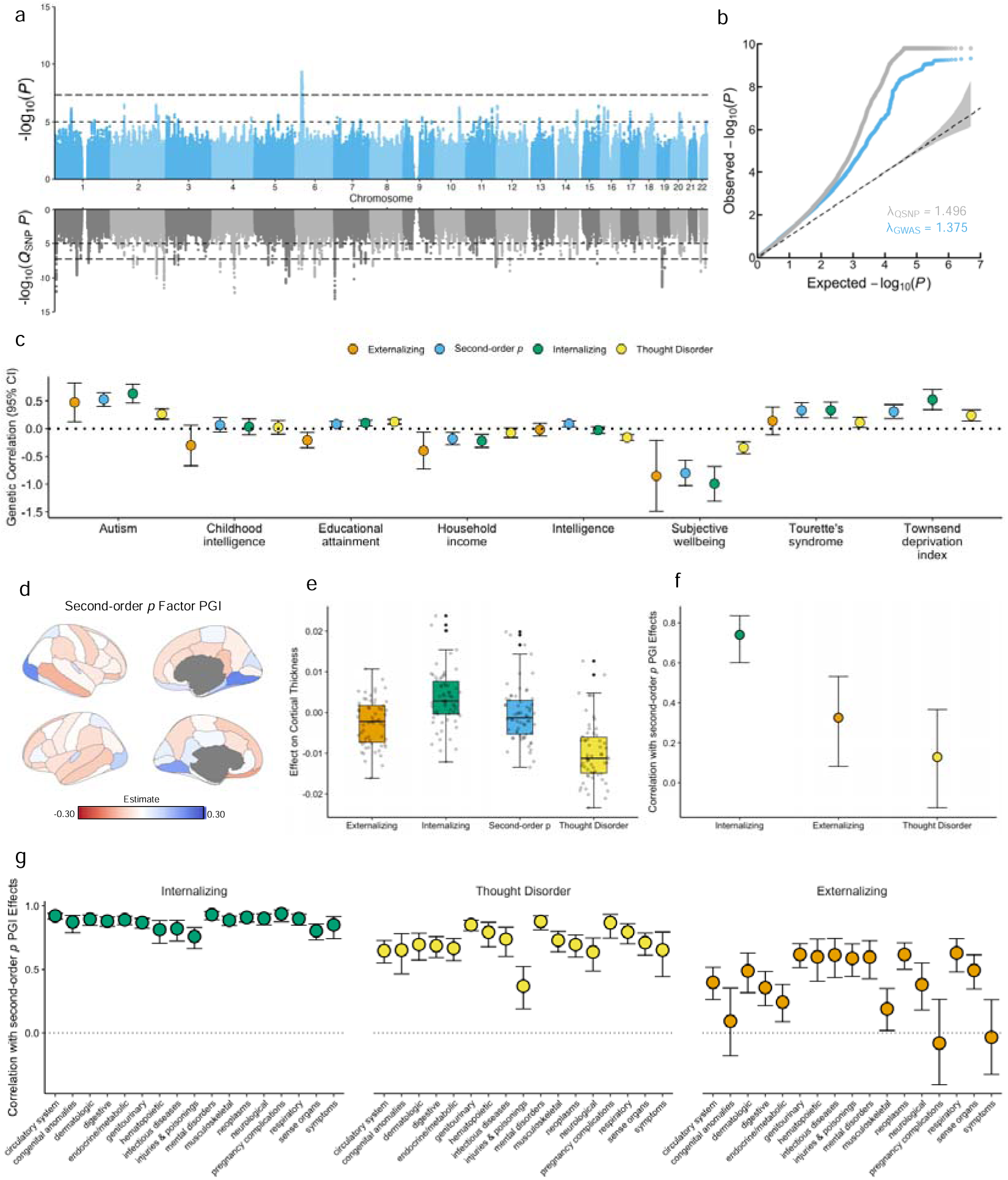
Comparison of *p* factor effects relative to domain-level factor effects. **a**, Miami plot for the second-order general psychopathology (*p)* factor. **b**, Q-Q plot of the second-order *p* factor and *Q*_SNP_ results. **c**, Scatter plot of genetic correlations between psychopathology factors and neurodevelopmental disorders, socio-economic outcomes, and subjective well-being. The dotted line indicates a genetic correlation of 0. **d**, Cortical map depicting associations between the second-order *p* factor polygenic index (PGI) and regional cortical thickness, as parcellated in the Desikan-Killiany-Tourville atlas. Effects were adjusted for age, sex, age^2^, and sex-by-age interactions (**Method**). **e**, Box plots depicting the distributions of PGI effects on regional cortical thickness. **f**, Scatter plot illustrating the correlation between standardized betas from the regression of the second-order *p* factor PGI on cortical thicknesses and those from the correlated factors model. **g**, Scatter plot of the correlations between the effect size of the associations of the *p* factor and the domain-level factors across phenome-wide association study outcomes in Mass General Brigham Biobank.

To evaluate whether the psychopathology factors were differentially related to risk for adverse medical outcomes, we conducted phenome-wide association study (PheWAS) analyses for each PGI in the Mass General Brigham Biobank (MGBB; *N* = 43,323). Once again, we found that the domain-level PGIs varied in their associations with health, genetic, and lifestyle outcomes (**Supplementary Fig. 25-27; Supplemental Information)**. However, unlike the cerebral results, the *p* factor PGIs were more broadly associated with health outcomes when compared to the individual domain-level PGIs. Effect sizes of the *p* factor PGIs were similar to one or more of the domain-level PGIs (Fig. 4g) – even for the 34 phecodes that were uniquely associated with *p*. Therefore, the *p* factor estimates generally resembled those of one or two specific factors, rather than all forms of psychopathology (**Supplementary Tables 14)**. Finally, despite the greater predictive power of the *p* factor PGIs, 9% of significant domain-specific associations were not observed when using these indices (**Supplemental Information)**.

## Discussion

Psychiatric disorders are a common cause of human suffering^3,43^, and their frequent co-occurrence complicates the effective assessment and treatment of these conditions^7^. Here, we examined several data-driven and theoretical models of shared etiology to help explain the pervasive comorbidity observed in this domain. We modeled the phenotypic and genetic factor structure of 10 psychiatric conditions in the UK Biobank to characterize patterns of (dis)similarity between these levels of analysis, which yielded novel insights into the form, validity, and utility of transdiagnostic factors in biological psychopathology. Three key insights emerged from these efforts.

First, the factor structure of mental conditions was only partially concordant across phenotypic and genetic levels of analysis, somewhat supporting Cheverud’s conjecture^44^ (or the “phenotypic null hypothesis”^45^ in the case of factor analysis), which states that phenotypic and genetic correlations between complex traits can be assumed to be highly similar^38^. While the structure of thought disorder and internalizing psychopathology was largely consistent, our results suggest that externalizing and compulsive disorders may be differentially related to other conditions at phenotypic and genetic levels. As disparities were not a consequence of heterogeneous data or low statistical power, differences might reflect environmental influences that cause phenotypic correlations to deviate from genetic ones. For instance, compulsive conditions were more strongly phenotypically (*r*= 0.889) than genetically (*r*= 0.449) correlated with internalizing conditions. Such deviations also occurred at the indicator level: OCD and AUD were phenotypically but not genetically correlated. One possible explanation is that overlapping symptoms may arise from a combination of different genetic predispositions and environmental factors. In contrast, the genetic correlation of depression and anxiety was greater (*r*= 0.908) than its phenotypic correlation (*r*= 0.618), which could suggest that while these disorders share genetic predispositions, environmental factors may drive different phenotypic expressions. These results provide new hypotheses to be tested by future gene-environment studies.

Second, we provide novel evidence for the validity and utility of multiple transdiagnostic factors in biological psychopathology, especially those characterized by internalizing, externalizing, compulsive, and thought disorders. With few exceptions, correlations between these factors were quite moderate, underscoring that they are largely modeling (co)variances that are unique from one another^5^. The internalizing factor, serving as an exemplar with the greatest number of indicators, was shown to be particularly robust to configural changes to the model. Moreover, despite some structural differences across levels of analysis, all four of these factors improved statistical power compared to the *p* factor GWAS and exhibited low rates of heterogeneity in genetic analyses, as demonstrated in the *Q*_SNP_ and *Q_Trait_* results.

Third, the *p* factor had relatively low validity and utility at the genetic level. While first- and second-order *p* factor models can be fit to the data, they offer limited insights into biology due to heterogeneous genetic effects across mental conditions. Specifically, *Q*_SNP_ and *Q_Trait_* results indicated that many SNP-level effects and nearly all genetic correlations were inconsistent with a general factor of psychopathology. Moreover, PGI results from the cerebral and PheWAS analyses revealed that information is lost when opting to use PGIs for *p* over correlated-but-distinct domain-level factors (e.g., thought disorder, internalizing, externalizing). Collectively, our findings build upon recent studies that suggest the *p* factor will generate fewer robust and comprehensive findings than the domain-level factors in the study of psychiatric comorbidity^46^ – at least with the currently available data.

Although the present study has taken important steps to address potential confounds and shortcomings, several limitations should be considered when interpreting results. For instance, the present study does not comprehensively sample the full spectrum of psychopathology and related conditions, as the UK Biobank did not have adequate data for all phenotypes of interest (e.g., autism spectrum disorder, attention-deficit/hyperactivity disorder, specific substance use disorders, schizoaffective disorder). This resulted in the omission of a neurodevelopmental factor^8,9^ and limited the available data for the thought disorder, externalizing, and compulsive factors, which were each modeled with two indicators. While these findings should be replicated in models with more data, our thought disorder and externalizing factors did have genetic correlations that were indistinguishable from unity (*r*_g_ = 1) with more expansive models from the literature^40,42^. Finally, our analyses were limited to individuals of European ancestries in the UK Biobank due to data availability and methodological constraints^25,36,47^. Extending these analyses to additional cohorts with more diverse data^48^ and less healthy volunteer bias^49^ will be critical to evaluating the robustness and generalizability of our findings.

In conclusion, the present study generates novel insights into the structure of psychopathology by reporting results at phenotypic *and* genetic levels, refining our understanding of comorbidity and shared etiology^10^. We add to the relatively scarce literature characterizing Cheverud’s conjecture and its exceptions in this domain, identifying instances where environmental influences may have a particularly pronounced effect on comorbidity. We also extend recent investigations into the validity of transdiagnostic factors in psychiatric genetic research^9,40,42,50,51^, providing practical insights that can guide the development and use of transdiagnostic PGIs in the field. Ultimately, our results highlight the intricate interplay between genetic and environmental factors in mental disorders^10,52^, paving the way for targeted studies of the shared etiological landscape for these conditions.

## Methods

### UK Biobank

The UK Biobank has phenotypic, genotypic, and imaging data from more than 500,000 participants. Participants were recruited between 2006 and 2010 across 22 assessment centers in England, Wales, and Scotland, between the ages of 40 to 69 years old^49^. Participants provided informed consent (https://biobank.ctsu.ox.ac.uk/crystal/field.cgi?id=200) and the UK Biobank received ethical approval from the Research Ethics Committee (reference 11/NW/0382). This study was conducted based on application 46007.

#### Genotyping, imputation, and quality checks

Participants (N = 488,377) were genotyped with the UK BiLEVE or the UK Biobank Axiom array. The array design, genotyping, and quality control procedures have been previously described by the UK Biobank investigators^53^. Genotypes were imputed to the Haplotype Reference Consortium (HRC) reference panel^26^ (Version 1.1) and UK10K and the 1000 Genomes Project Phase 3 reference panel by the UK Biobank. We kept SNPs from the 1000 Genomes Project Phase 1^54^ that were filtered on minor allele frequency ≥ 0.01, genotyping call rate ≥ 0.05, and a Hardy-Weinberg equilibrium threshold ≥ 5×10–6. Samples were filtered on missingness rate ≥ 0.1. In total, 5,319,661 variants and 487,409 people pass filters and QC, including 431,006 individuals from British ancestry with genetic principal components.

#### Image acquisition and preparation

We analyzed the cortical mean thicknesses and surface areas of the Desikan-Killiany-Tourville (DKT) atlas from the first Magnetic Imaging Resonance (MRI) visit generated by an image-processing pipeline developed and run by the UK Biobank Imaging team (Category 196^55,56^). Analyses were conducted in R^57^. The MRI data were collected with a standard Siemens Skyra 3 T running VD13A SP4 with a standard Siemens 32-channel RF receive head coil. The UK Biobank Imaging team analyzed the 3D MPRAGE T1-weighted volumes with pipeline scripts that primarily call for FSL and Freesurfer tools. The UK Biobank Imaging Protocols provide details of the acquisition protocols, image processing pipeline, image data files, and derived measures of brain structure and function. There were three scanner sites located in Cheadle (Site 11025, coded as 0); Reading (Site 11026, coded as 2); and Newcastle (Site 11027, coded as 3).

#### Psychiatric phenotype construction

We used 10 binary lifetime psychiatric disorders or conditions diagnoses (depression (DEP), generalized anxiety (ANX), suicidality (SUI), eating disorders (EAT), posttraumatic stress disorder (PTSD), obsessive-compulsive disorders (OCD), Hazardous Alcohol Use or Dependence (AUD), substance use disorder (SUD), bipolar disorder(BIP), and schizophrenia (SCZ)) generated in a previous study^58^ by combining self-reported measures and patient diagnoses (**Supplementary Table 1**). We did not include psychiatric disorders or conditions in the UK Biobank with fewer than 1,000 cases, such as autism spectrum disorder and attention-deficit/hyperactivity disorder. If a participant reported having a psychiatric disorder or condition during any of their visits at the center or the online follow-up, they were identified as a case for that disorder. There were 502,120 individuals with data on the presence or absence of a disorder for at least 1 of the 10 disorders (501,234 without missing data) and cases ranged from 1,3210 (OCD) to 101,530 (depression; **Table 1**; **Supplementary Table 2**). These analyses were conducted in R^57^ and were not restricted to British individuals.

#### UK Biobank holdout sample

We created a validation holdout sample of UK Biobank individuals with neuroimaging data that were not related to individuals in the UK Biobank discovery sample. This yielded 24,162 individuals (12,511 females) for which we calculated polygenic indices.

### Factor analyses

We calculated the phenotypic and genetic correlations across psychiatric disorders and conducted exploratory factor analyses to identify models of psychopathology to validate in the confirmatory factor analyses.

#### Bivariate correlations among study phenotypes

We calculated the bivariate phenotypic and genetic correlations (*r*_p_ and *r*_g_) for all study phenotypes to better understand patterns of comorbidity and shared etiology among psychiatric disorders. For the phenotypic data, we randomly selected 50% of the sample as the training sample and calculated the tetrachoric correlation matrix of our psychiatric conditions using the *mixedCor* function from the *psych* package^59^ in R^57^. The phenotypic correlation here assesses the degree to which two psychiatric conditions tend to vary together in a population. For the genetic data, we used LD Score Regression^25^ to calculate the genetic correlation from all SNPs. A high genetic correlation suggests that there is a strong genetic overlap between the two traits, indicating that the same genes are likely involved in the expression of both traits.

#### Disparity calculations

To quantify differences between the phenotypic and genetic correlations of the 10 psychiatric conditions, we calculated the ‘disparity’^38,39^ between bivariate phenotypic and genetic correlations (*D* = *r*_p_ – *r*_g_) in R^57^. To evaluate whether the calculated disparities were systematically related to technical factors, we computed Pearson correlations between *D* estimates and (1) the standard errors of genetic correlations, (2) the proportion of external cases in contributing GWAS, and (3) the effective sample size of contributing GWAS. For the latter two tests, we used a permutation-based correlation approach. Specifically, for each disorder, we shuffled the vector values during each of the 1,000 permutations and then computed the correlation between the matrix and this permuted vector. This yielded a distribution of correlations under the null hypothesis of no association specific to each disorder. The observed correlation for each disorder was then compared to its respective null distribution to compute empirical *p*-values. When these *p*-values are non-significant, it suggests that observed discrepancies might reflect environmental influences leading to differences in phenotypic expression.

#### Phenotypic factor analyses

We ran exploratory factor analyses (EFA) on the tetrachoric correlation matrix of our psychiatric conditions from the training sample using the *fa* function from the *psych* package^59^ in R^57^. Since indicators were categorical and some loadings were under 0.5, we used the difference in the Root Mean Square Error Approximation (ΔRMSEA) with a 0.015 cut-value to determine the number of factors to retain^60^. We first examined the ΔRMSEA across EFAs with 1-4 factors. We applied an oblimin rotation and the principal axis factoring (PAF) method, which is more appropriate for non-normal data than the maximum likelihood method^61^. We report results on the sample with missingness for some psychiatric conditions because the model fit and variance explained across psychiatric disorders or conditions were similar when excluding individuals with missing data.

We used the *lavaan* package ^62^ in R^57^ to conduct confirmatory factor analyses (CFA) in the test sample (i.e., the remaining 50% of the sample) with and without missing data using diagonally weighted least squares (DWLS) estimation and pairwise likelihood estimation for missing cases (Fig. 1). CFA models were informed by the published literature^23^, as well as the EFA results. We ran bifactor models with modified latent variable variances and loadings and added constraints when the solution was not found. Specifically, we constrained the loadings on residual latent factors with only 2 indicators to solve Heywood cases^63^.

Good fit was assessed with the following cut-offs: a comparative fit index (CFI) > 0.95, a root mean square error approximation (RMSEA) < 0.06, and a standardized root mean square residual (SRMR) < 0.08 (Hu and Bentler 1999). We report the model fit and parameters from the models with participants that had missing data or one or more disorders since the model parameters were similar when including and excluding individuals with missing data.

#### Genetic factor analyses

To characterize the genetic structure of psychopathology, we first performed EFA with promax rotation and modeled up to 4 factors in the training data, a genetic covariance matrix constructed from odd chromosome SNPs. We then used Genomic SEM^36^ to evaluate the fit of these data-driven models in the test data in R^57^, a genetic covariance matrix constructed from even chromosome SNPs, before testing model fit on all SNPs. Finally, we also used Genomic SEM to estimate SNP effects on the latent genetic factors. Estimated sample size (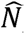) for each factor was calculated as the mean of 1/((2 * MAF * (1 - MAF)) * SE^2) across SNPs^42^.

### Genome-wide association analyses

#### Univariate genome-wide association analyses

We ran GWASs of the 10 psychiatric conditions in the UK Biobank (**Supplementary Table 1**) in 395,364 individuals of white British ancestry who did not have neuroimaging data. UK Biobank GWASs were conducted using a sparse GRM (fastGWA-GLMM^64^) from the *Genome-wide Complex Trait Analysis (GCTA)* package^65^. We controlled for relatedness, sex (0 = males, 1 = females), center, genotyping chip, birth year, and the first 40 PCs of the genotyped data. We included information provided by the UK Biobank on the genetically inferred kinship of respondents (estimated in the KING software^66^).

#### Meta-analysis

Since several psychiatric conditions had few cases in the UK Biobank, we sought to improve statistical power of our GWASs via meta-analysis. For this procedure, we obtained GWAS summary statistics for similar phenotypes (**Supplementary Table 1**) that (i) were publicly available, (ii) excluded the UK Biobank, and (iii) were from European ancestry. We then meta-analyzed the UKB and external summary statistics using METAL^67^. Meta-analyzed GWAS summary statistics (META GWASs) were generated for all disorders except for SUD, which lacked publicly available summary statistics due to the broad definition of case status. Meta-analyses were conducted using a sample size weighted analysis. The weight was set to the effective *N* for univariate summary statistics (Neff = 4*(sample.prevalence*(1-sample.prevalence))*(N controls + N cases)) and to the sum of the N Effective (i.e., the sum of the Neff of each cohort included in the meta-analysis) for meta-analyzed summary statistics^68^. When the minor allele frequency (MAF) was unavailable, we calculated the MAF from the reported frequency of the effect allele or used the MAF from the 1000 Genome Project Phase 3 reference panel^54^.

#### Multivariate genome-wide association analyses

Prior to analysis, all summary statistics were munged using Genomic SEM, where we applied the conventional filters of imputation quality ≥ 0.90 and minor allele frequency (MAF) ≥ 0.01. The genetic covariance and sampling covariance matrices were estimated using multivariable LDSC in Genomic SEM^36^. Model fit was assessed using the following cut-offs: comparative fit index (CFI) > 0.95 and Standardized Root Mean Square Residual (SRMR) < 0.08 indicative of good fit, and CFI > 0.90 indicative of acceptable fit. Diagonally-weighted least squares estimation was used in all Genomic SEM analyses.

To characterize the genetic structure of psychopathology, we first performed EFA with promax rotation and modeled up to 4 factors in the training data, a genetic covariance matrix constructed from odd chromosome SNPs. We then used Genomic SEM^36^ to evaluate the fit of these data-driven models in the test data, a genetic covariance matrix constructed from even chromosome SNPs, before testing model fit on all SNPs. Finally, we also used Genomic SEM to estimate SNP effects on the latent genetic factors. Estimated sample size (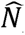) for each factor was calculated as the mean of 1/((2 * MAF * (1 - MAF)) * SE^2) across SNPs^42^.

We used the “functional mapping and annotation of genetic associations” method (FUMA; version 1.3.5e^69^) to extract (i) the number of lead SNPs and genomic loci associated with a factor and (ii) the number of lead SNPs and genomic loci identified in the *Q*_SNP_ analyses (see the section below) using the multivariate GWAS results from Genomic SEM as input. We then compared the number of lead SNPs from the first-order factor analyses to those from the *Q*_SNP_ analyses (we set the maximum cutoff *P* threshold to 1). The remaining parameters were set to their default value (see SNP2GENE: https://fuma.ctglab.nl/tutorial~snp2gene). FUMA was applied on about 5,319,661 SNPs for each multivariate GWAS output.

We performed *Q*_SNP_ analyses to examine whether a SNP operates through a given factor. This metric is described elsewhere^36,70,71^. In brief, it is an index of the decrement in model fit between two models: one where the factor of interest is regressed onto a given SNP and another where the indicators for the factor of interest are regressed onto a given SNP or trait. Subsequent comparison of these two models allows us to test whether the observed pattern of association plausibly operates via the latent factor or if it is better described by indicator-specific relationships. We calculated the factor and the *Q*_SNP_ mean χ^2^ on the reference SNPs from the 1000 Genomes Project Phase 3 reference panel^54^ as the mean of the squared Z estimate obtained for each SNP from the Genomic SEM output.

### Comparing the Phenotypic and Genetic Psychopathology Factor Models

We conducted three sets of analyses to compare phenotypic and genetic factor structure of our phenotypes. First, we examined the consistency of the UK Biobank and META GWAS results to ensure that differences in factor model structure were not due to idiosyncrasies between the genetic and phenotypic data that were introduced by including public GWAS results. This was accomplished by (i) examining differences in the genetic correlation and the heritability of the indicators from the UK Biobank and META GWASs using LDSC^25^, (ii) conducting 2-, 3-, and 4-factor EFAs on the UK Biobank and the META GWAS results, (iii) estimating the correlation of the indicator loadings of the UK Biobank GWAS and META GWAS across genetic and phenotypic models, and (iv) examining whether the correlation between the meta-analyzed genetic correlation matrix was more similar to the phenotypic correlation matrix than the UK Biobank genetic correlation matrix. Second, we tested the phenotypic model in the genetic data and vice-versa to test whether the models fit similarly across data types. Third, when applied to the genetic and phenotypic data, we calculated the correlation of factor loadings across the genetic and phenotypic models.

### Genetic correlations with external traits

We examined the concurrent validity of our psychopathology factors by estimating genetic correlations between our factors and factors from models in the literature with additional indicators and larger samples. Specifically, we examined the correlation between our externalizing factor and the externalizing factor from a well-powered multivariate GWAS excluding 23andMe data^40,41,^. We also estimated the genetic correlation between our internalizing factor and a factor indexing mood disturbance (self-reported depressive, psychotic, and manic symptoms, as well as clinically diagnosed bipolar II and major depressive disorder), as well as the genetic correlation between our thought disorder factor and a factor indexing serious mental illness (bipolar I, schizoaffective disorder, and schizophrenia). We recreated the latent factors from the literature using the model structure and indicator GWAS from the literature and calculated the genetic correlation between the latent factors from the literature and ours with Genomic SEM^36^. Our latent factors were estimated with our indicator GWAS.

We performed *Q*_Trait_ analyses to examine whether the pattern of genetic correlation between an outcome ad the indicators of the model are well accounted for by the latent factor. The calculation of this statistics is described elsewhere^36,70,71^. In brief, it is an index of the decrement in model fit between a model where the factor of interest is regressed onto a given trait and a model where the indicators for the factor of interest are regressed onto a given trait. We compare these models to test whether the observed pattern of association plausibly operates via the latent factor or if it is better described by indicator-specific relationships.

For the *Q*_Trait_ analyses, we used GWAS summary statistics from a previously published pipeline^40,42,68^, while removing phenotypes that were redundant with the investigated latent factor (e.g., schizophrenia was excluded from the pipeline when estimating genetic correlations with the thought disorder factor). Summary statistics were chosen due to their broad relevance to human health and well-being, as previously described^40,42^. Briefly, they correspond to 4 main domains: health and disease outcomes, personality and risky behavior, psychopathology and cognition, and demography and socioeconomic status. The number of external traits for which *Q*_Trait_ analyses were conducted depended on whether the *Q*_Trait_ model converged. In some cases, the *Q*_Trait_ model did not converge because of negative variance estimates.

### Polygenic index (PGI) analyses

#### PGI construction

Using sBayesR^72^, we created polygenic indexes (PGIs) for individuals with neuroimaging data and their siblings with the summary statistics of the latent variables of the most parsimonious and best-fitting genetic and phenotypic models.

#### PGI analyses in UK Biobank

We created residualized PGIs by adjusting the PGIs for the year of birth, genetic sex, and the first 40 principal components of the genotyped data. We excluded individuals with related individuals in the discovery and individuals that were related in the target sample. The major histocompatibility complex (MHC) region was excluded from the PGI analyses for the COMP and first-order *p* factor PGI because including the MHC region led to counterintuitive results (i.e., first-order *p* factor PGI predicted over 3% of the TD factor but 0.8% of the first-order *p* factor). Removing the MHC region did not affect the PGI prediction of the other psychopathology factors.

To examine the predictive power of the PGIs on their respective latent variables, we quantified the degree to which each PGI predicted the latent psychopathology factors in the present study (Fig 3.). The loadings were set to those reported in the behavioral analyses (Fig. 3a**,c**) CFA test model to address Heywood cases and model convergence issues. The residualized PGI was the regression predictor and each latent factor served as an outcome. We fit the complete measurement model per PGI, with each PGI predicting each factor (i.e., the INT PGI predicting EXT, TD, and INT in the same model). We report the *R*^2^ of each PGI for all eight psychopathology factors. The following cutoffs were set to establish a good model fit: CFI > 0.95, RMSEA < 0.06, and SRMR < 0.08^73^.

To evaluate associations between the PGIs of psychopathology and cortical brain measures, we examined the association between the PGIs and 64 regional mean thicknesses and 64 surface areas from the cortical Desikan-Killiany-Trouville atlas. We conducted linear regressions with PGIs, age at the MRI, sex, quadratic age at imaging, sex and age interactions, and sex and quadratic age interactions predicting each cortical measure. Age corresponds to age at MRI and the PGI were adjusted for year of birth and the 40 first genetic principal components provided by the UK biobank. Continuous variables were unit standardized to have a mean of 0 and a standard deviation of 1. We report the correlations between the brain-PGI associations to compare (1) brain-PGI associations between the *p* factors and domain-specific factors and (2) brain-PGI associations between domain-specific factors.

#### PGI analyses in MGB Biobank

To evaluate associations between the PGIs of psychopathology and a wide variety of medical outcomes, we performed PheWAS analyses in the Mass General Brigham Biobank (MGBB). The MGBB is a biorepository from the MGB healthcare system with patient data on electronic health record, genetic, and lifestyle variables^74^. It has enrolled 138,042 participants, 65,265 of whom have also been genotyped. The recruitment strategy, genotyping procedures, and quality control procedures for this dataset have been described in previously published studies^74^. All participants provided written consent upon enrollment, and analyses were conducted under MGB Institutional Review Board protocols #2009P002312 and #2021P003641.

A standard “phecode” approach^75,76^ was used to assign case status for 1,817 medical outcomes, requiring the presence of at least two International Classification of Disease 10 Clinical Modification (ICD-10-CM) codes. To reduce the risk of population stratification, PheWAS analyses were restricted to 43,323 patients of European ancestry. PGIs were computed using the sBayesR method^64^ in these individuals, as described above, and standardized prior to analysis. PheWAS analyses were then conducted using the PheWAS package in R^77^ (https://github.com/PheWAS/PheWAS), where logistic regression models were fit for each of the 1,817 medical outcomes under study. Sex assigned at birth, current age, genotyping chip, and the first 10 genetic principal components were included as covariates. Statistical significance was evaluated at a Bonferroni-corrected threshold (*p* < 2.75e-5).

To evaluate whether there was any sample overlap between our GWAS and PheWAS data, we used LD Score regression to estimate the cross-trait intercept between a published GWAS of tobacco use disorder in the MGBB^43^ and our correlated factors of psychopathology. The genetic correlation estimates were non-zero in all cases (EXT *r*_g_ = 0.592 [*SE* = 0.120], INT *r*_g_ = 0.210 [*SE* = 0.066], and TD *r*_g_ = 0.171 [*SE* = 0.059]), which implies the cross-trait intercept can be used to detect sample overlap. We found the cross-trait intercept was not significantly different from zero in all cases, indicating that there is no detectable overlap between the GWAS and PheWAS data in the present study.

## Data Availability

The data is accessible to researchers with an ongoing application in the UK Biobank.

https://osf.io/unkym/?view_only=eea9ecdd47324c65b0e9b494925a178

## Acknowledgments

Funding for investigator effort has been provided by Agence Nationale de la Recherche (ANR-17-EURE-0017 and ANR-10-IDEX-0001-02 PSL) to F.R. TTM is supported by funds from NIH T32HG010464.

A CC-BY public copyright license has been applied by the authors to the present document and will be applied to all subsequent versions up to the Author Accepted Manuscript arising from this submission, in accordance with the grant’s open access conditions.

## Data/code availability

The data is accessible to researchers with an ongoing application in the UK Biobank. Multivariate GWAS results and the study’s code are accessible here: https://osf.io/unkym/?view_only=eea9ecdd47324c65b0e9b494925a1783.

