## Supplemental Files for "Characterizing the phenotypic and genetic structure of psychopathology in UK Biobank"

Supplemental Information

### **Results**

#### 1. **Comparing the UK Biobank Genome-Wide Association Study (GWAS) results to the Meta-analyzed (META) GWAS results**

To ensure that discrepancies between the phenotypic and genetic factor models were not due to sampling differences we used the UK Biobank phenotypic and genetic data. However, given that the UK Biobank had too few cases for some psychiatric conditions, we included external cohorts. To confirm that discrepancies between our phenotypic and genetic psychopathology models did not stem from sampling differences, we ensured that the UK Biobank GWAS results were similar to those from the META GWAS.

We found discrepancies for psychiatric phenotypes that substantially gained power following the meta-analyses (i.e., SCZ) or that still had low power after the meta-analysis (e.g., EAT). However, the genetic correlations and heritability (**Supplementary Fig.1**), the exploratory factor analysis (EFA) structure (**Supplementary Fig.2**), and the factor loadings from the confirmatory factor analysis (CFA) (**Supplementary Fig.3**) were generally consistent across UK Biobank and META GWAS results.

In more detail, the heritability estimates were generally unchanged and increased after including public GWAS results for some phenotypes with relatively few cases in the UK Biobank, including SCZ, BIP, and EAT (**Supplementary Fig.1**). The genetic correlations of the indicators from the UK Biobank and the meta-analyzed GWASs were also generally consistent, although they differed for some of the phenotypes with lower statistical power. Specifically, the genetic correlations of SCZ, BIP, and EAT with other indicators differed between the UK Biobank and meta-analyzed GWASs (median |∆| = 0.22, 0.16, and 0.16, respectively), which is unsurprising given that SCZ, BIP, and EAT had the most substantive increases in sample size via meta-analysis (**Supplementary Fig.1; Table 1**).

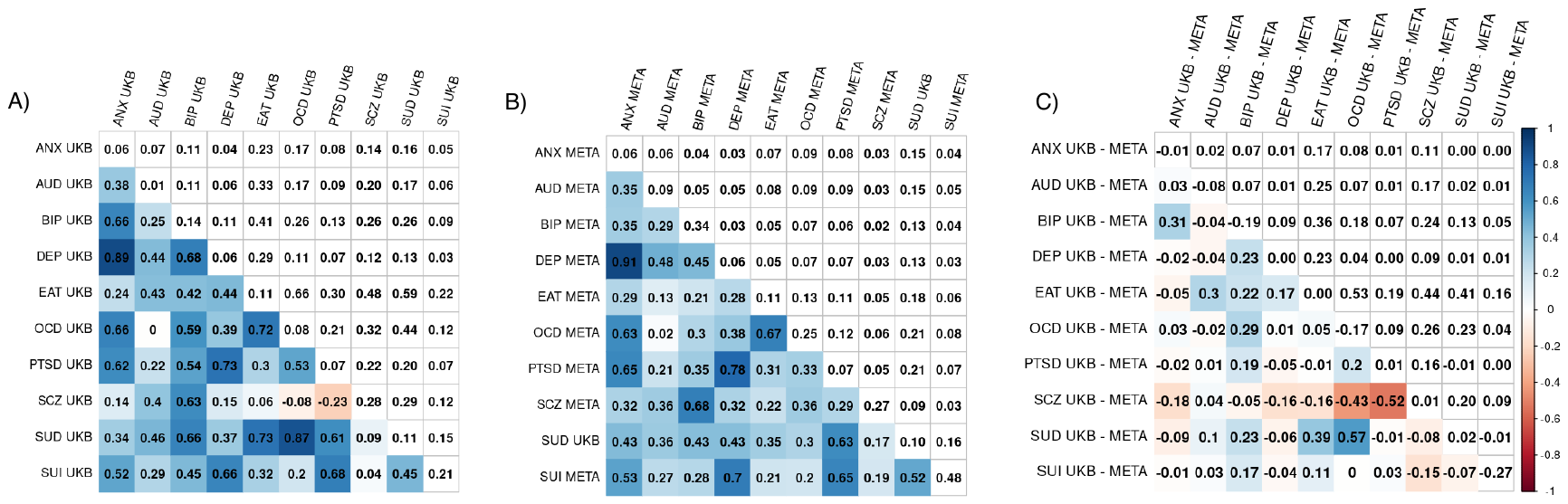

**Supplementary Fig.1. Genetic correlations and heritability estimates of 10 psychiatric conditions from (A) the UK Biobank and (B) the meta-analyzed results, and (C) their difference**. Lower and upper triangles display pairwise LD Score genetic correlation estimates and their standard errors, respectively and the diagonal displays observed-scale SNP heritability estimates (h2). Statistics from LD Score regression^1^. UKB (UK Biobank). Meta-analyzed (META). Alcohol use disorder (AUD), generalized anxiety (ANX), bipolar disorder (BIP), depression (DEP), substance use disorder (SUD*), eating disorder (EAT), obsessive-compulsive disorder (OCD), posttraumatic stress disorder (PTSD), schizophrenia (SCZ), and suicidality (SUI).

When conducting EFAs on all SNPs from the UK Biobank and the META GWAS results, we found that the factor structure was generally similar across GWAS results, except for the AUD and SUD indicators (**Supplementary Fig.2**).

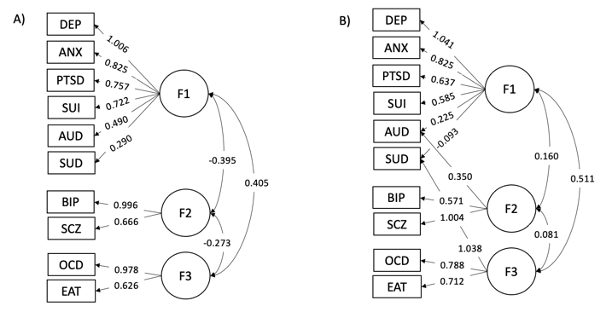

**Supplementary Fig.2. Exploratory factor analysis results on all SNPs from (A) the meta-analyzed and (B) the UK Biobank results**. Squares correspond to indicators, circles to latent variables, double-headed arrows to correlations, and single-headed arrows to factor loadings. Note that the factor loadings are regression coefficients and not correlations because the factors are correlated. For clarity, we only show regression coefficients over 0.200 and coefficients in panel (B) that are present in panel (A). Alcohol use disorder (AUD), generalized anxiety (ANX), bipolar disorder (BIP), depression (DEP), substance use disorder (SUD), eating disorder (EAT), obsessive-compulsive disorder (OCD), posttraumatic stress disorder (PTSD), schizophrenia (SCZ), and suicidality (SUI).

Indicator factor loadings were highly correlated across UK Biobank and META GWAS results for both the genetically and phenotypically informed models (**Supplementary Fig.3**). SCZ was the only indicator whose loadings changed between the UK Biobank GWAS and META GWAS results. We posit that this discrepancy might stem from the substantial increase in sample size in the meta-analysis.

Despite some discrepancies for the phenotypes that either showed significant improvement in statistical power following meta-analysis or continued to exhibit low power, the genetic correlations, heritability estimates, and factor analysis were largely consistent between the UK Biobank and the META GWAS results.

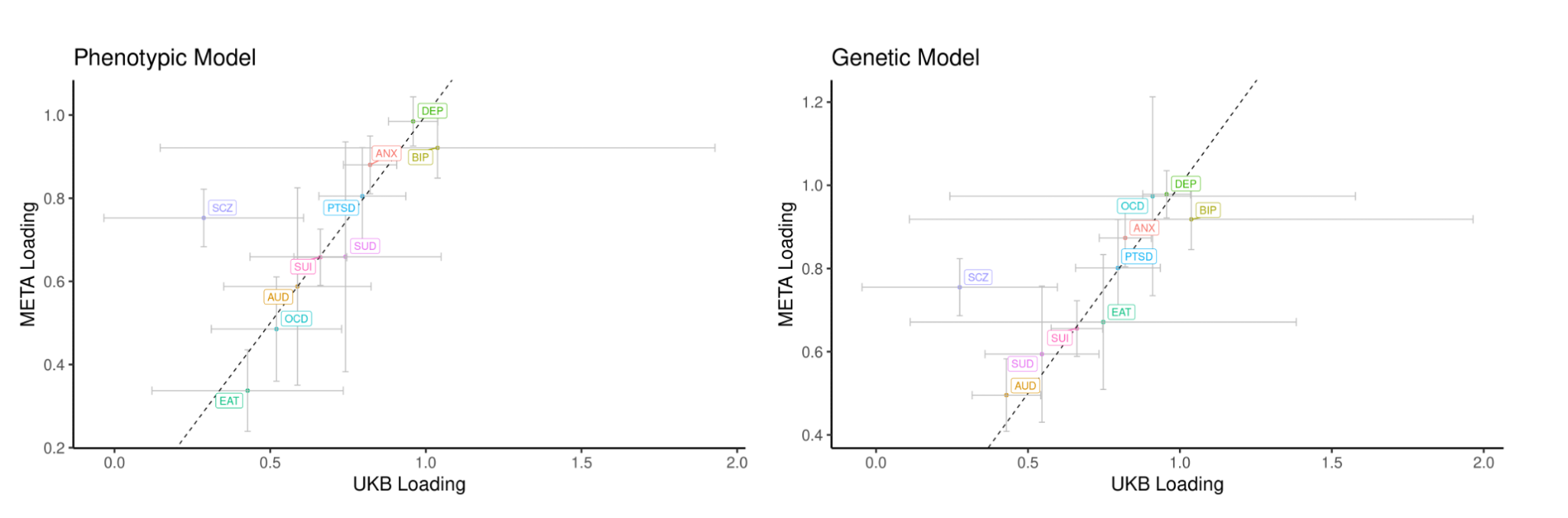

**Supplementary Fig.3. Correlation between factor loadings in the UK Biobank and the meta-analyzed data across the phenotypically and genetically informed models**. See Fig.5 for genetically and phenotypically informed models on the phenotypic data. Error bars reflect 95% confidence intervals calculated from the standard errors of the standardized estimate from genomic SEM^2^. Loadings refer to the standardized estimate from Genomic SEM. Alcohol use disorder (AUD), generalized anxiety (ANX), bipolar disorder (BIP), depression (DEP), substance use disorder (SUD), eating disorder (EAT), obsessive-compulsive disorder (OCD), posttraumatic stress disorder (PTSD), schizophrenia (SCZ), and suicidality (SUI).

#### 2. **Exploratory Factor Analysis**

To identify the number of factors to include in the exploratory factor analyses, we conducted a parallel analysis and examined a scree plot. The parallel analysis and the scree plot suggested that up to 5 factors be extracted from the data (**Supplementary Fig.4.a**). The difference in root mean square error of approximation (RMSEA) between the 3 and 4-factor solutions was under 0.15 (**Supplementary Fig.4.b**) indicating that a 3-factor solution sufficiently describes the data. Loadings for each genetic and phenotypic factor solution are available in **Supplementary Table 5**.

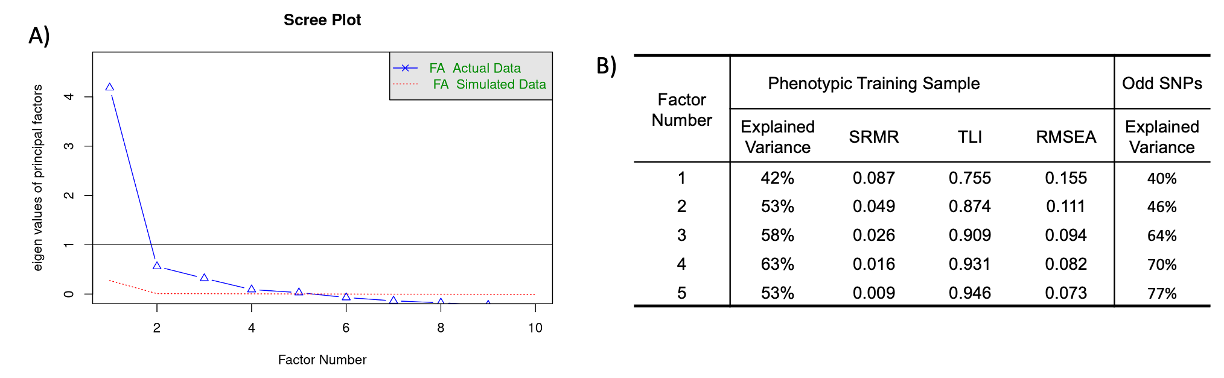

**Supplementary Fig.4. Exploratory factor analysis (EFA) results.** Panel A corresponds to a scree plot, indicating that 5 factors or less are sufficient to describe the data. Panel B is a table of the explained variance and model fit of EFAs with 1 to 5 factors. Factor Analysis (FA). Root Mean Square Error of Approximation (RMSEA). Standardized Root Mean Squared Residual (SRMR). Tucker-Lewis Index (TLI). Single Nucleotide Polymorphisms (SNPs).

#### 3. **Genetic and Phenotypic Psychopathology Models**

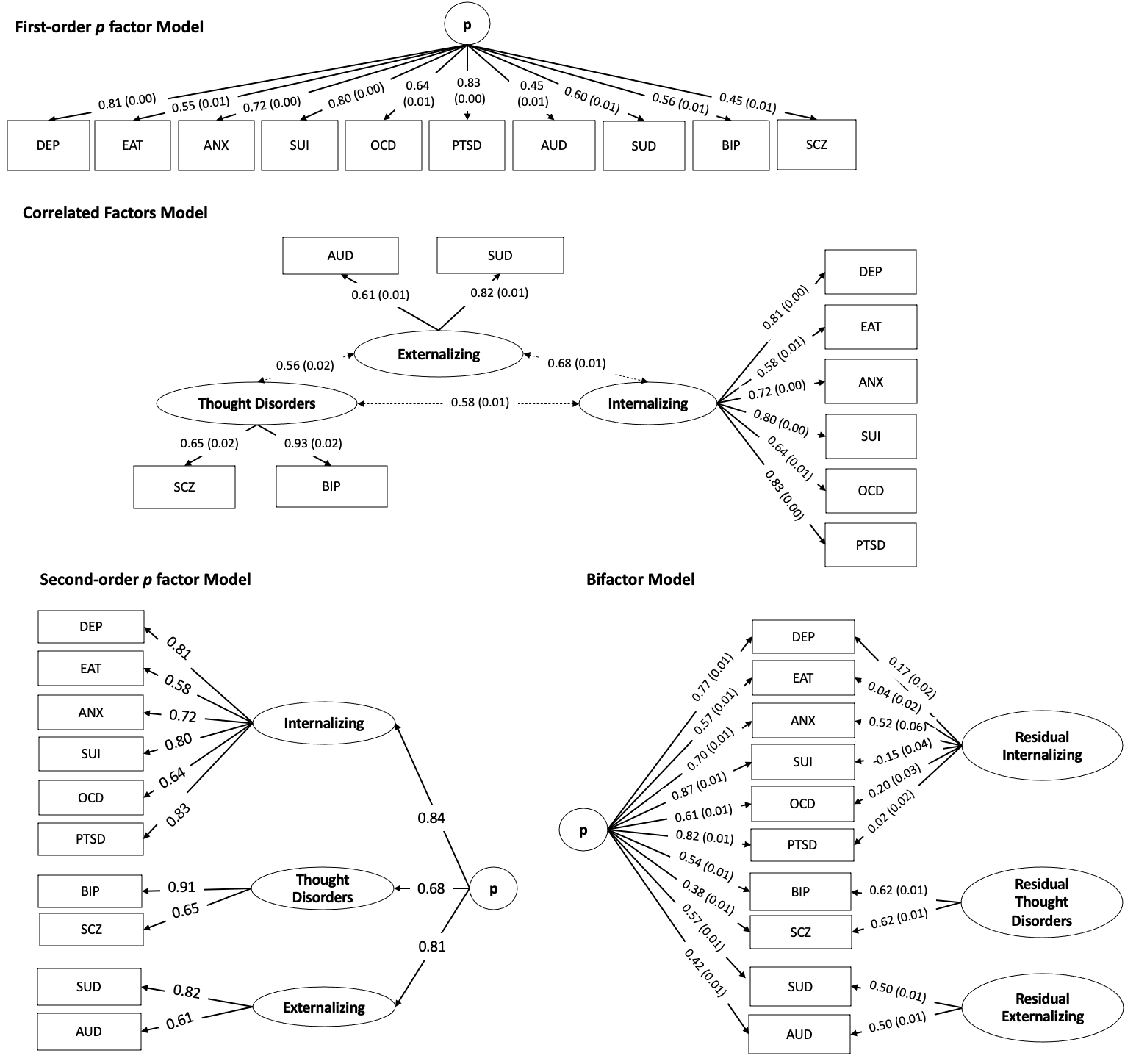

#### **Supplementary Fig.5. Phenotypic psychopathology models applied to the phenotypic data.** Squares correspond to indicators, circles to latent variables, double-headed arrows to correlations, and single-headed arrows to factor loadings. For simplicity, we report indicator and factor variance in **Supplementary Table 6**. Alcohol use disorder (AUD), generalized anxiety (ANX), bipolar disorder (BIP), depression (DEP), substance use disorder (SUD), eating disorder (EAT), obsessive-compulsive disorder (OCD), posttraumatic stress disorder (PTSD), schizophrenia (SCZ), and suicidality (SUI).

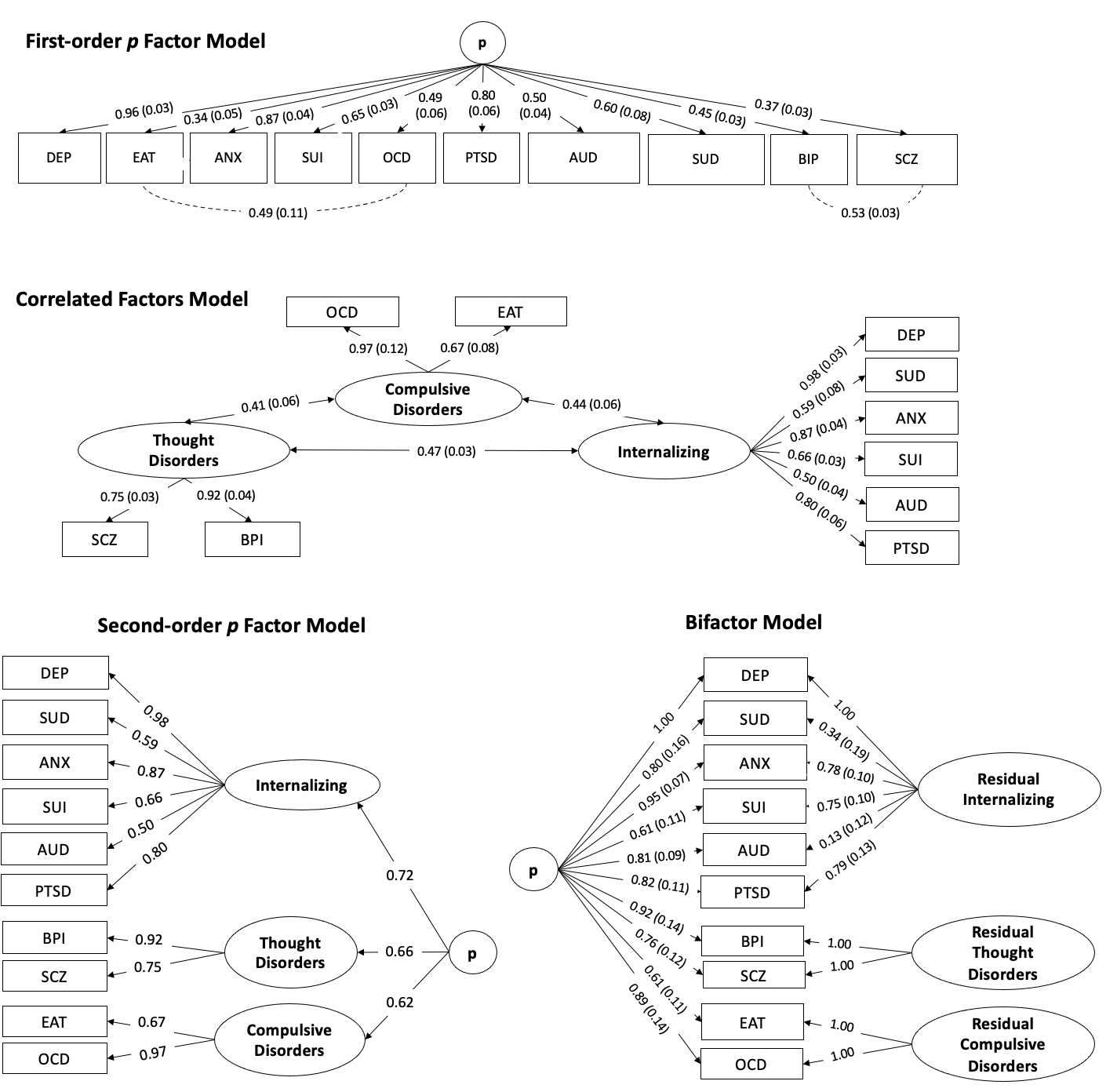

#### **Supplementary Fig.6. Genetic psychopathology models applied to the genetic data.** Squares correspond to indicators, circles to latent variables, double-headed arrows to correlations, and single-headed arrows to factor loadings. For simplicity, we report indicator and factor variance in **Supplementary Table 8**. Alcohol use disorder (AUD), generalized anxiety (ANX), bipolar disorder (BIP), depression (DEP), substance use disorder (SUD), eating disorder (EAT), obsessive-compulsive disorder (OCD), posttraumatic stress disorder (PTSD), schizophrenia (SCZ), and suicidality (SUI).

#### 4. **Sensitivity Analyses on the Internalizing Factor**

We performed a series of analyses to characterize the concordance between the internalizing factor from the phenotypically informed model (INT_p_) and the internalizing factor from the genetically informed model (INT_g_).

We used Genomic SEM^39^ to estimate the SNP effects of the transdiagnostic factors from the phenotypic and genetic correlated factors model. We calculated the mean χ^2^ restricted to the SNPs in the high-quality consensus genotype set defined by the HapMap 3 Consortium ^64^ and estimated the genetic correlation between INT_p_ and INT_g_. The mean χ^2^ of the INT_p_ and INT_g_ GWAS were both 1.45, using the 678,838 SNPs in the high-quality consensus defined by the HapMap3 Consortium (**Table 2**). They were also genetically correlated at 0.99, suggesting that they index the same genetic signal (**Supplementary Fig. 19 and 21** for Manhattan and Miami plots, respectively).

Using the “functional mapping and annotation of genetic associations” method (FUMA^65^, **Methods**), we identified the number of lead SNPs and genomic loci associated with both factors and examined their genetic overlap at the SNP level. We found that 25 lead SNPs and 19 genomic loci were associated with INT_p_ and that 24 lead SNPs and 18 genomic loci were associated with the INT_g_ (**Table 2**). There were 5 genomic loci associated with INT_p_ that were not associated with INT_g_. Third, we ran *Q*_SNP_ analyses to examine whether the same SNPs operated through the INT_p_ and INT_g_ factors. There were no *Q_SNP_* hits for INT_p_ or INT_g_, suggesting that all significant SNPs operate through the Internalizing factor.

Using LD score regression implemented in Genomic SEM^2^, we examined the genetic correlation of INT_p_ and INT_g_ with 68 external traits. The pattern of genetic correlations was highly similar between INT_p_ and INT_g_ (**Supplementary Fig.7**, **Supplementary Table 15**), suggesting that the convergent and discriminant validity of the INT factor is not impacted by configural changes between INT_p_ and INT_g_.

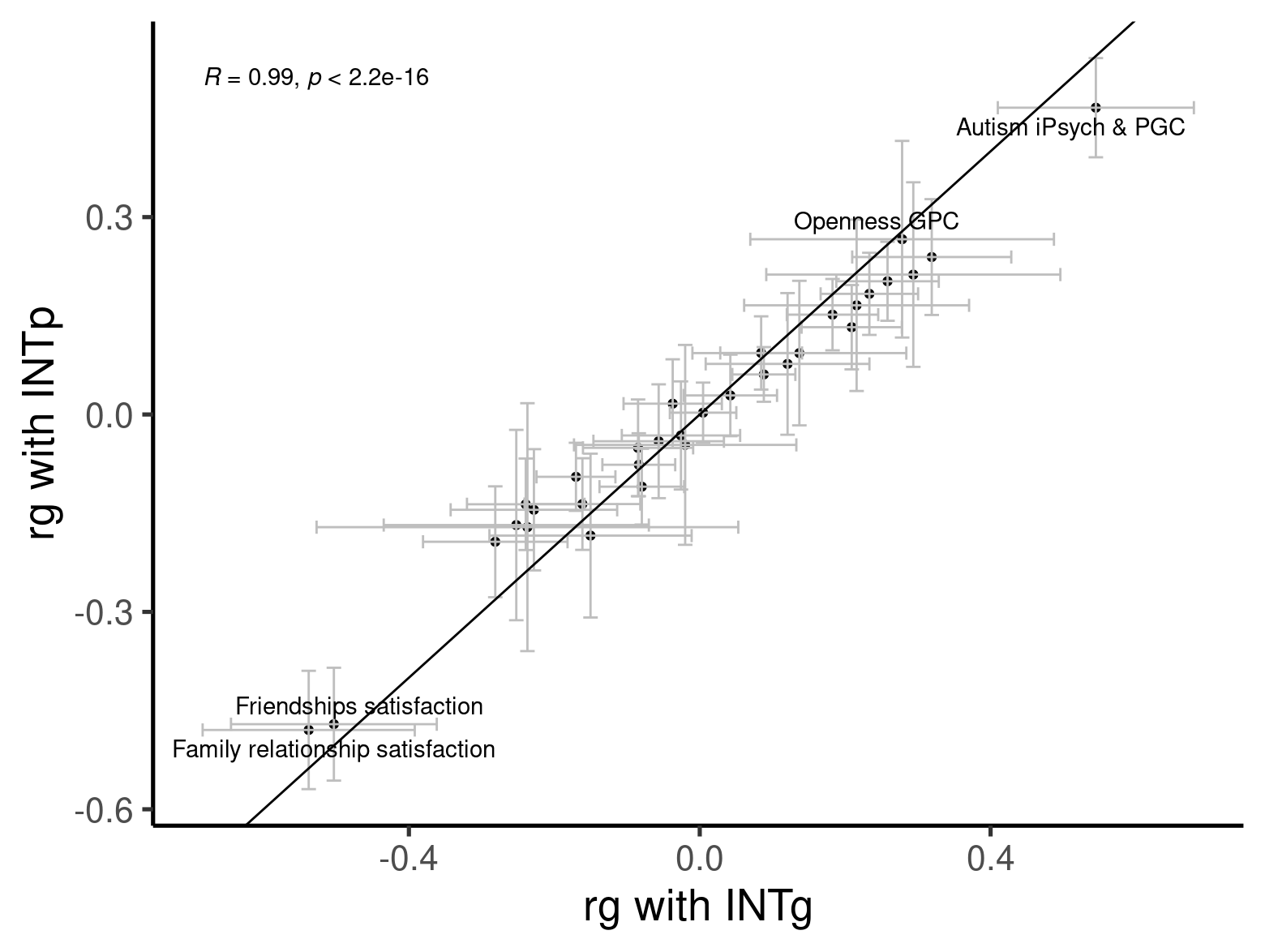

**Supplementary Fig.7. Genetic correlation (rg) between the loadings of the internalizing factor (INT) from the UK Biobank and the meta-analyzed genome-wide association studies across genotypic (INTg) and phenotypic (INTp) models**. Error bars reflect 95% confidence intervals calculated from the standard errors of the standardized estimate from Genomic Structural Equation Modelling (SEM). Loadings refer to the standardized estimate from Genomic SEM.

We then examined whether the polygenic indices (PGI s) based on INT_p_ and INT_g_ differed in the predictive power. We found that both the INT_p_ and INT_g_ polygenic indices explained about 3% of internalizing in our UK Biobank holdout sample. Finally, we examined the convergence in the polygenic index-brain association between the INT_p_ and INT_g_ polygenic indices and found that the association between the polygenic index and cortical volumes, mean thicknesses, and surface areas from the Desikan-Killiany-Tourville parcellation were highly correlated across INT polygenic indices (r = 0.998; **Supplementary Fig.8**).

Finally, we conducted a PheWAS for each PGI in genotyped individuals of European ancestry in the Mass General Brigham Biobank (MGBB) biorepository with a logistic regression fit to 1,819 case/control disease phenotype. We found that the effects of the internalizing PGI on disease phenotypes were correlated at 0.985 (95%CI[0.9823, 0.986]) between the INT_p_ and INT_g_ PGIs.

Considered together, these analyses suggest that the internalizing factor is robust across configurations: the same latent trait is captured when AUD and SUD are replaced with OCD and EAT.

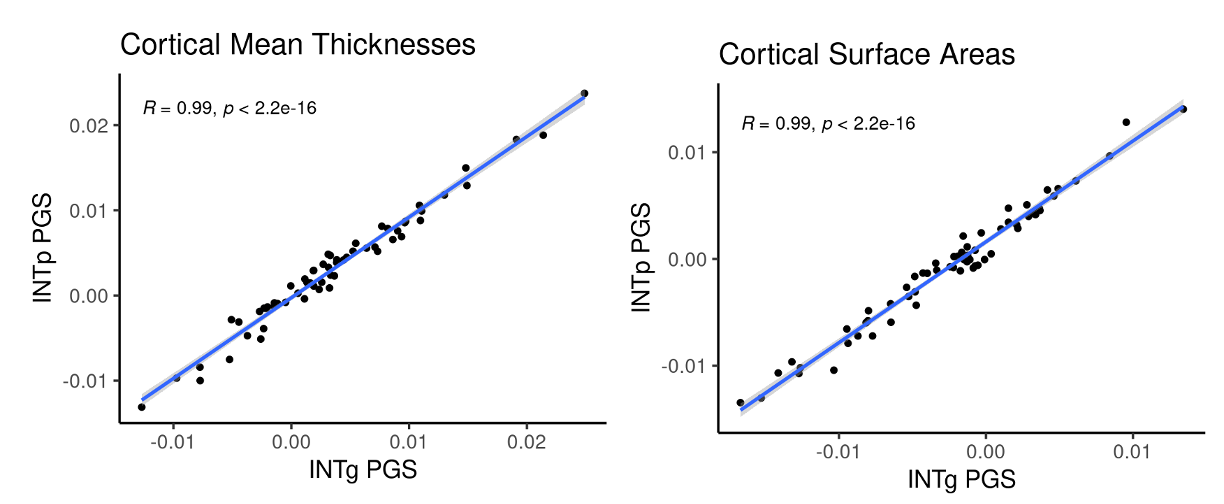

**Supplementary Fig.8. Correlations between the brain-polygenic index association of the internalizing polygenic index (PGI) from the phenotypically-informed model (INT_p_) and the internalizing PGI from the genetically-informed model (INT_g_).** Cortical mean thicknesses and surface areas from the Desikan-Killiany-Tourville parcellation. Pearson correlation (R). Points: standardized estimate of polygenic index-region association. Blue regression line.

To further examine the internalizing factor's robustness, we re-estimated the correlated factors model from the phenotypic model with a reduced number of indicators, which we refer to as the reduced INT models. We refer to the original internalizing factor from the phenotypic model as the full internalizing model (i.e., INT = DEP + OCD + PTSD + EAT + ANX + SUI). We correlated the loadings of the reduced INT models with those from the full INT models to evaluate whether the factor loadings differed between the original and reduced models. We focused on reduced models with 3 indicators. We tested models with OCD, EAT, and each internalizing indicator to examine whether adding an indicator to OCD and EAT yielded a similar latent factor to the internalizing one obtained in the full model. We then performed a series of other analyses where we include two internalizing indicators and either OCD or EAT (i.e., INT = DEP + OCD + PTSD and INT = EAT + ANX + SUI) to examine whether two internalizing indicators in addition to OCD or EAT are sufficient to yield a similar latent factor to the internalizing one obtained in the full model.

We found that the loadings from the full model differed from the loadings of the reduced model when the reduced model included OCD and EAT and another internalizing indicator. This suggests that the latent factor from the reduced model differs from that of the full model. However, when additional internalzing indicators are added in the model (e.g., the internalizing factor includes OCD, EAT, PTSD and DEP) or when the reduced model include OCD or EAT and two other indicators, we find similar loadings (**Supplementary Fig.9**), suggesting that the latent factor across these reduced models and the full models are consistent. We note that while the Comparative Fit Index (CFI) indicated a good model fit, the Standardized Root Mean Squared Residual (SRMR) indicated an acceptable model fit (< 0.10).

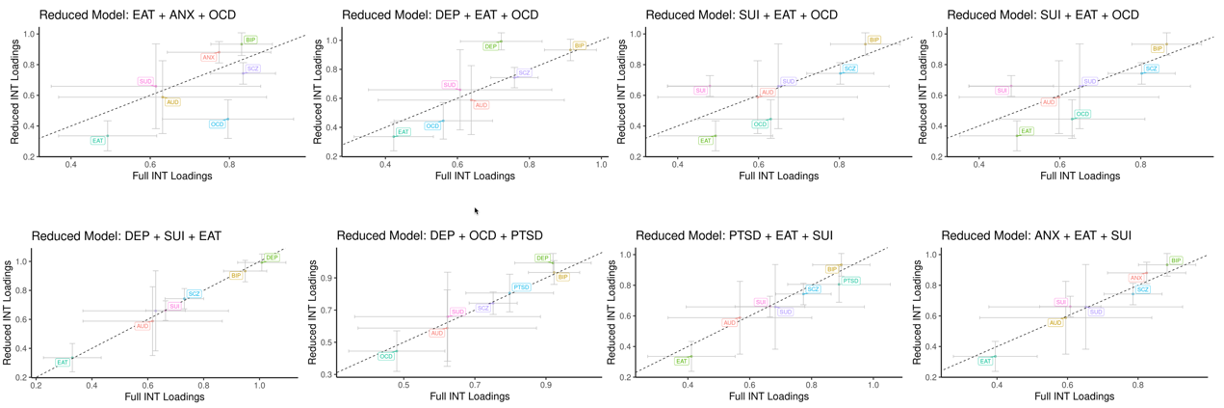

**Supplementary Fig.9. Correlations among factor loadings for internalizing models with different indicators.** Full internalizing (INT) model includes generalized anxiety (ANX), depression (DEP), eating disorder (EAT), obsessive-compulsive disorder (OCD), posttraumatic stress disorder (PTSD), and suicidality (SUI). Slope of dashed black line = 1 and error bars the 95%CIs.

#### 5. **The bifactor model applied to the genetic data**

We focused on the first- and second-order *p* factor from the phenotypic first- and second-order *p* factor models respectively because the bifactor model did not fit the genetic data well: The bifactor p-factor model fit was acceptable (CFI = 0.982, SRMR = 0.079), although it did not converge unless the first indicator loading of a factor was set to unity. Moreover, the residual variance of depression was negative. Constraining the residual variance of depression above 0 solved the Heywood case but lead to another warning, indicating model instability. Although constraining the residual variance of depression to be above 0 had little effect on the model parameters, the loadings of the indicators on p in the bifactor differed from those of the first-order p-factor model. When estimating SNP effects, we obtained additional warnings regarding a non-positive definite covariance matrix. The model did not converge when specifying an alternate bifactor structure where the subfactors were correlated.

#### 6. ***Q_SNP_* & *Q_Trait_* Analyses**

We conducted *Q_SNP_* and *Q_Trait_* analyses to examine the heterogeneity of the psychopathology factors. The *Q_SNP_* and *Q_Trait_* analyses showed high levels of heterogeneity for the second-order *p* factor. In the *Q_SNP_* analyses, 35 genomic loci were *Q* hits and 1 genomic locus was a factor hit, indicating that 35 genomic loci did not solely operate through the p-factor. Only one genomic locus was associated with the *p* factor (**Supplementary Fig.10**). The high degree of heterogeneity (*Q_SNP_* mean χ^2^ = 1.475) across the 3 factors of the second-order *p* factor GWAS may explain why there was only one factor hit even though the association mean χ^2^ of the *p* factor was high (1.505; **Table 2**). In the *Q_Trait_* analyses of 73 external traits, 31 out of the 38 trait factor hits (82%) were also *Q* hits, indicating that the patterns of genetic associations between external traits and each domain-level factor were not well accounted for by the second-order *p* factor (**Supplementary Fig.11**).

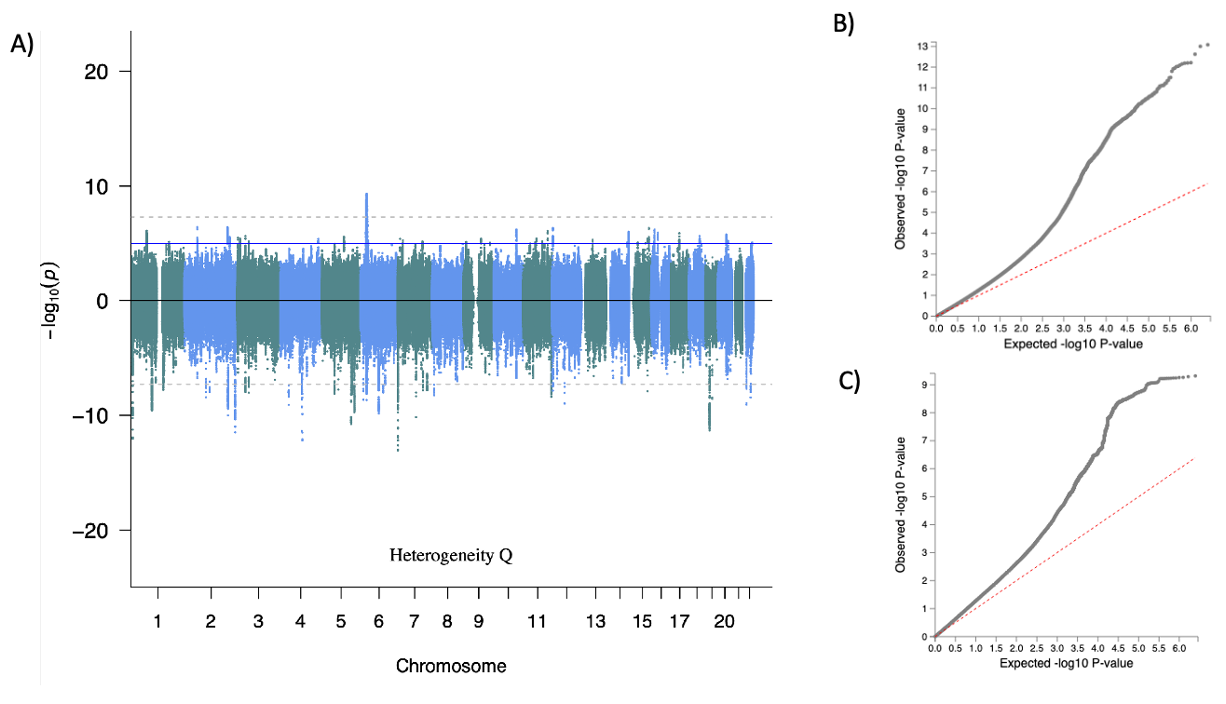

**Supplementary Fig.10. Miami plot (A), factor Q-Q plot (B), and *Q*_SNP_ Q-Q plot (C) from the phenotypically informed second-order *p* factor** **model*.*** Dotted grey "genome-wide significant" line at -log10(5e-8) and blue "suggestive" line at -log10(1e-5). Panel (A) top: first-order factor Genome-wide Association Study (GWAS) results. Panel (A) bottom: heterogeneity *Q* GWAS.

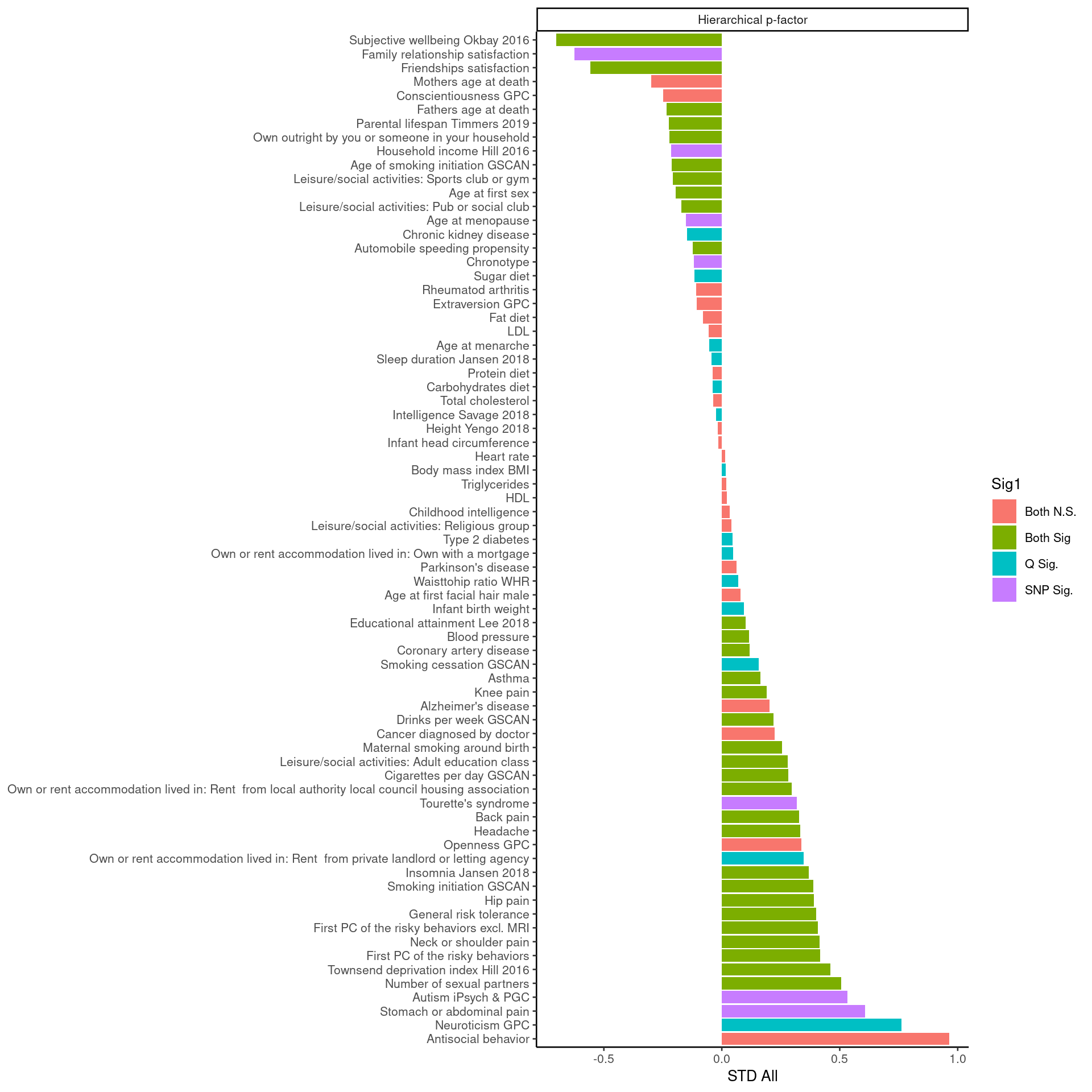

**Supplementary Fig.11. Q_Trait_** **analyses for the second-order p factor.** The significance threshold is set at p < 0.05/73 (the number of external traits). Both N.S. (no factor or *Q* hits). Both Sig. (Significant factor and *Q* hit). *Q* Sig. (Significant *Q* hit, but not factor hit). SNP Sig. (Significant factor hit, but not *Q* hit). A significant *Q* hit indicates that the pattern of genetic correlations between the indicators and the external trait is not well accounted for by the factor.

More genomic loci were associated with the p-factor from the first-order p-factor model compared to the p-factor from the second-order model: 47 genomic loci were associated with the p-factor, 57 genomic loci did not solely operate via the p-factor and only 6 were both factor and Q hits (**Table 2, Supplementary Fig.12**). However, the mean χ^2^ of the *Q_SNP_* analyses was greater than the mean χ^2^ of the first-order factor, suggesting greater heterogeneity in the genetic signal of the p-factor than polygenic architecture. Therefore, the *Q_SNP_* hits suggest heterogeneity in the p-factor. In the *Q_Trait_* analyses of 65 external traits, 36 out of the 38 factor hits (95%) were also *Q* hits, indicating that the patterns of genetic associations between external traits and each subfactor were not well accounted for by the first-order p-factor (**Supplementary Fig.13**). The second-order and first-order p-factor models were highly correlated (0.98; **Supplementary Fig.14**).

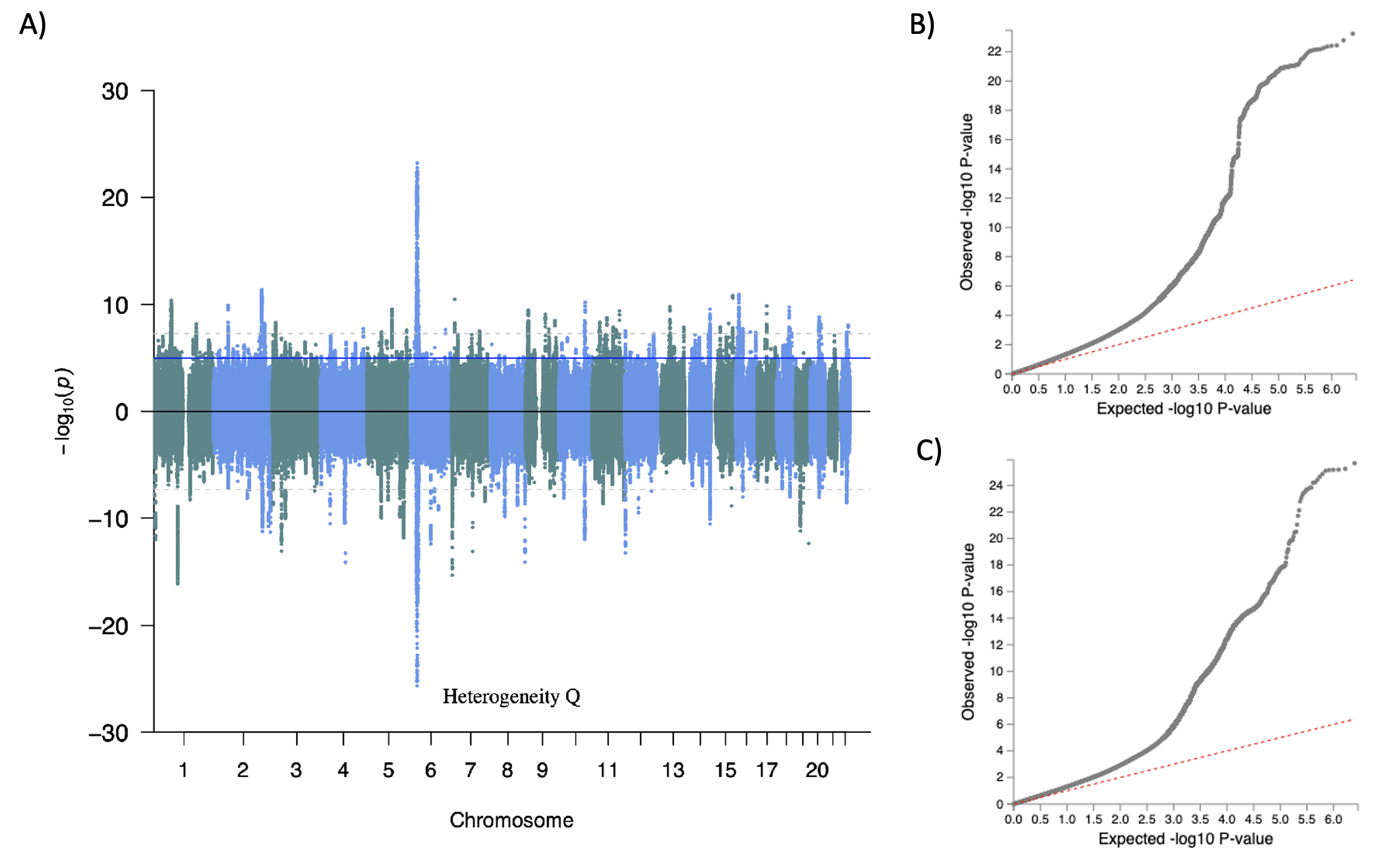

**Supplementary Fig.12. Miami plot (A), factor Q-Q plot (B), and *Q*_SNP_** **Q-Q plot (C) from the first-order *p* factor model.** Dotted grey "genome-wide significant" line at -log10(5e-8) and blue "suggestive" line at -log10(1e-5). Panel (A) top: first-order factor Genome-wide Association Study (GWAS) results. Panel (A) bottom: heterogeneity *Q* GWAS.

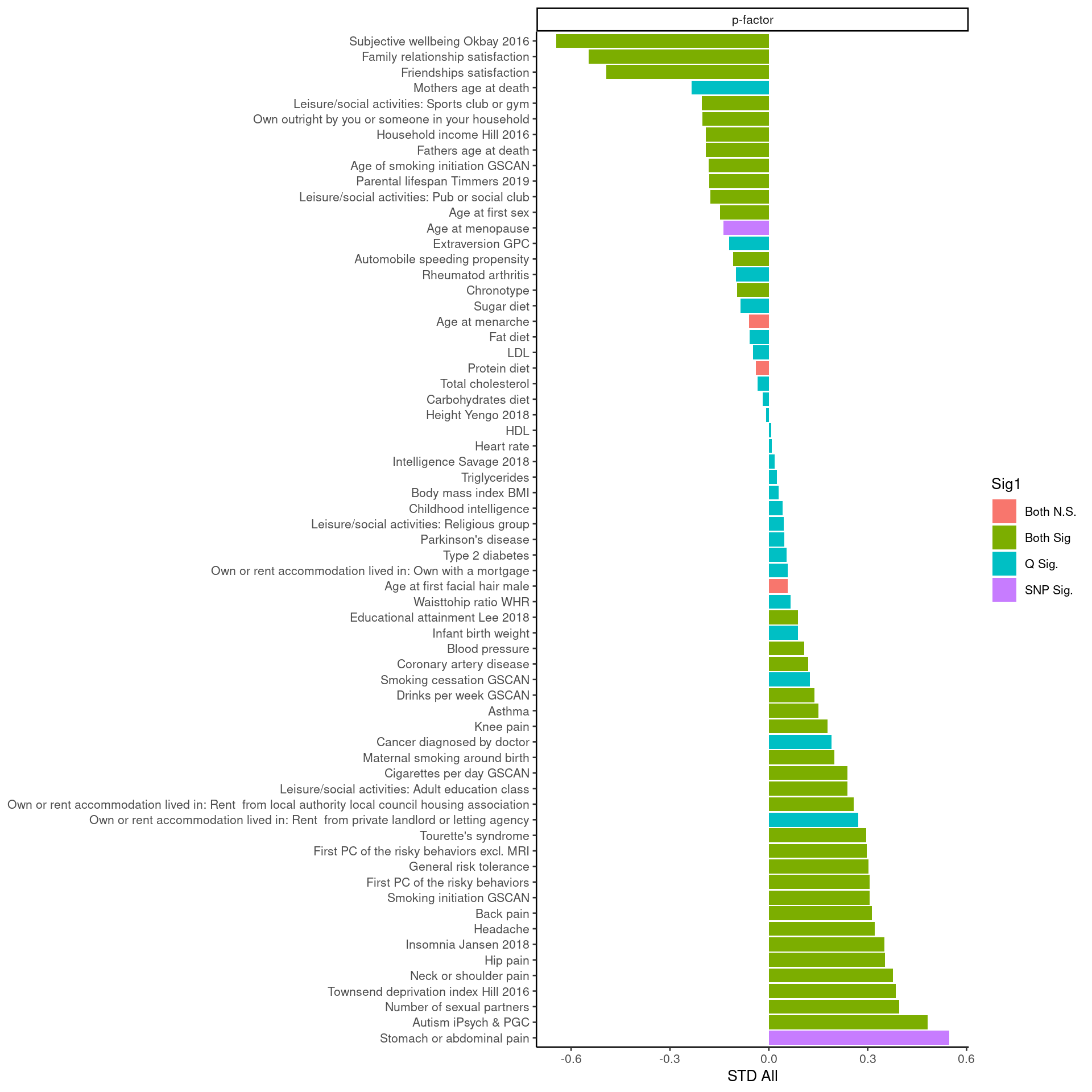

**Supplementary Fig.13. *Q_Trait_* analyses for the first-order *p* factor from the phenotypic model.** The significance threshold is set at p < 0.05/65 (the number of external traits). Both N.S. (no factor or *Q* hits). Both Sig. (Significant factor and *Q* hit). *Q* Sig. (Significant *Q* hit, but not factor hit). SNP Sig. (Significant factor hit, but not *Q* hit). A significant *Q* hit indicates that the pattern of genetic correlations between the indicators and the external trait is not well accounted for by the factor.

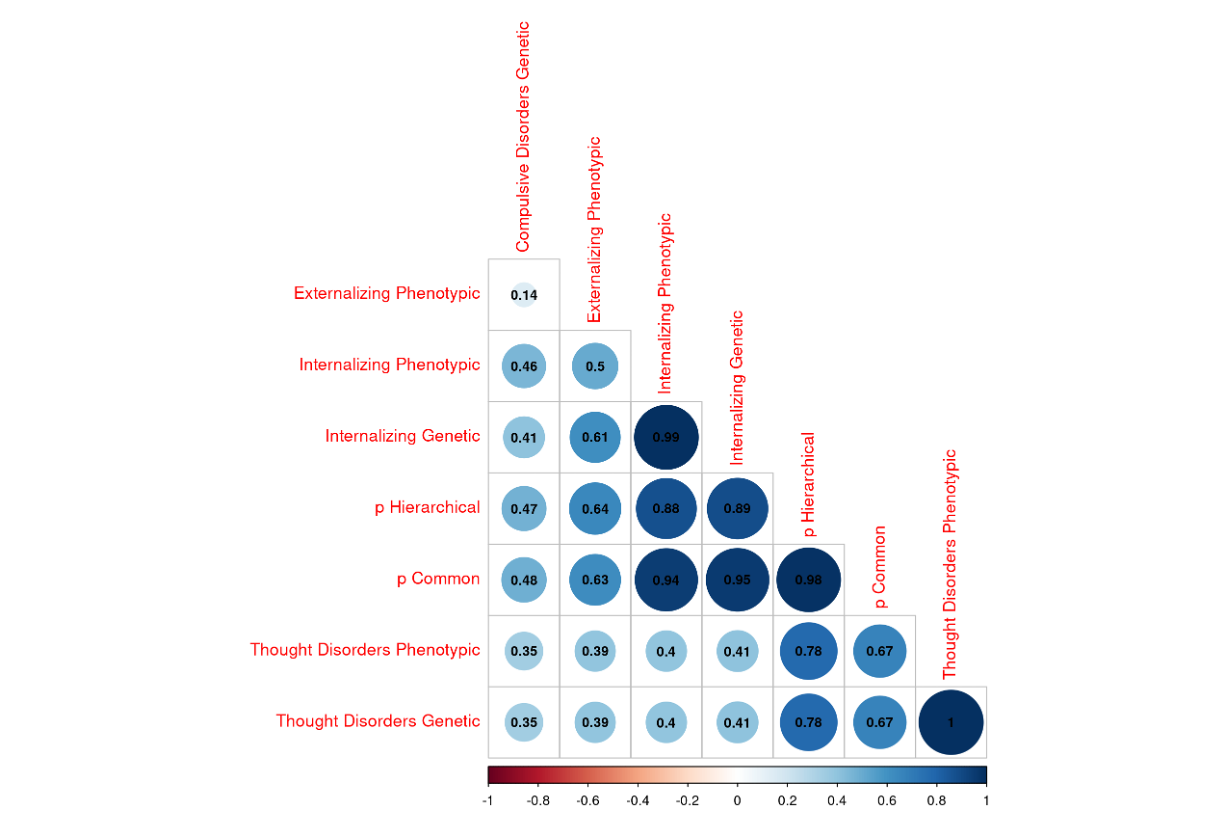

**Supplementary Fig.14. Genetic correlation between factors from the correlated factors genetic and phenotypic model, the *p* factor from the second-order model, and the *p* factor from the first-order *p* factor.**

As a point of comparison, we ran *Q_Trait_* and *Q_SNP_* analyses on the factors from the phenotypic correlated factors model:

The *Q_SNP_* analyses had 97-factor hits and 2 Q hits (**Table 2; Supplementary Fig.15**), suggesting that more SNPs operated through the thought disorders factors than SCZ and BIP. In the *Q_Trait_* analyses of 68 external traits on the thought disorders factor, 8 out of the 20-factor hits (40%) were also *Q* hits (**Supplementary Fig.16**), suggesting that the pattern of associations between the individual disorders and the external traits is well accounted for by the thought disorders factor. Therefore, the thought disorders factor appears to be biologically meaningful at the SNP level.

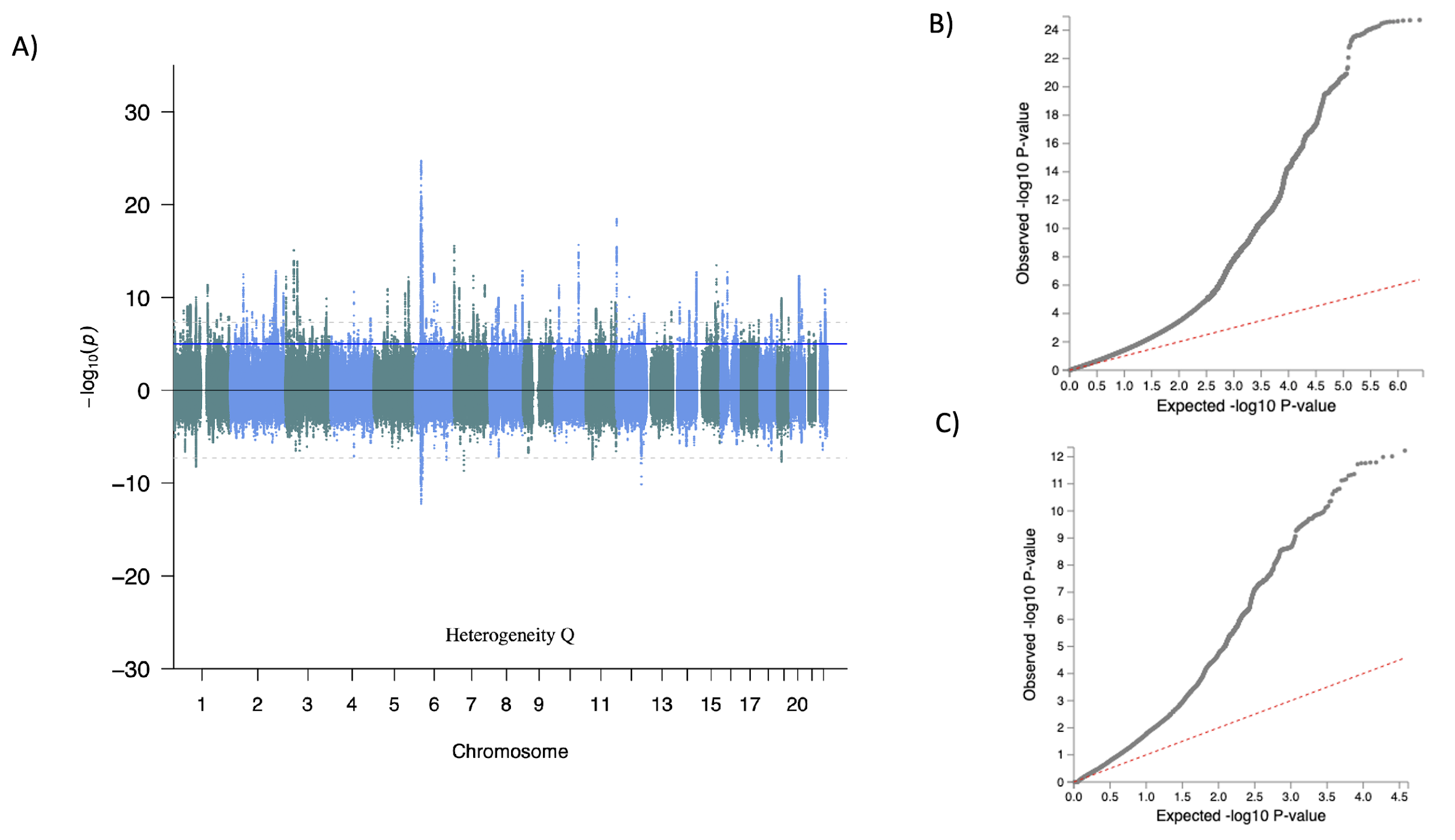

**Supplementary Fig.15. Miami plot (A), factor QQ-plot (B), and *Q*_SNP_** **QQ-plot (C)y of the thought disorders factor from the phenotypic correlated factors model.** Dotted grey "genome-wide significant" line at -log10(5e-8) and blue "suggestive" line at -log10(1e-5). Panel (A) top: first-order factor Genome-wide Association Study (GWAS) results. Panel (A) bottom: heterogeneity *Q* GWAS.

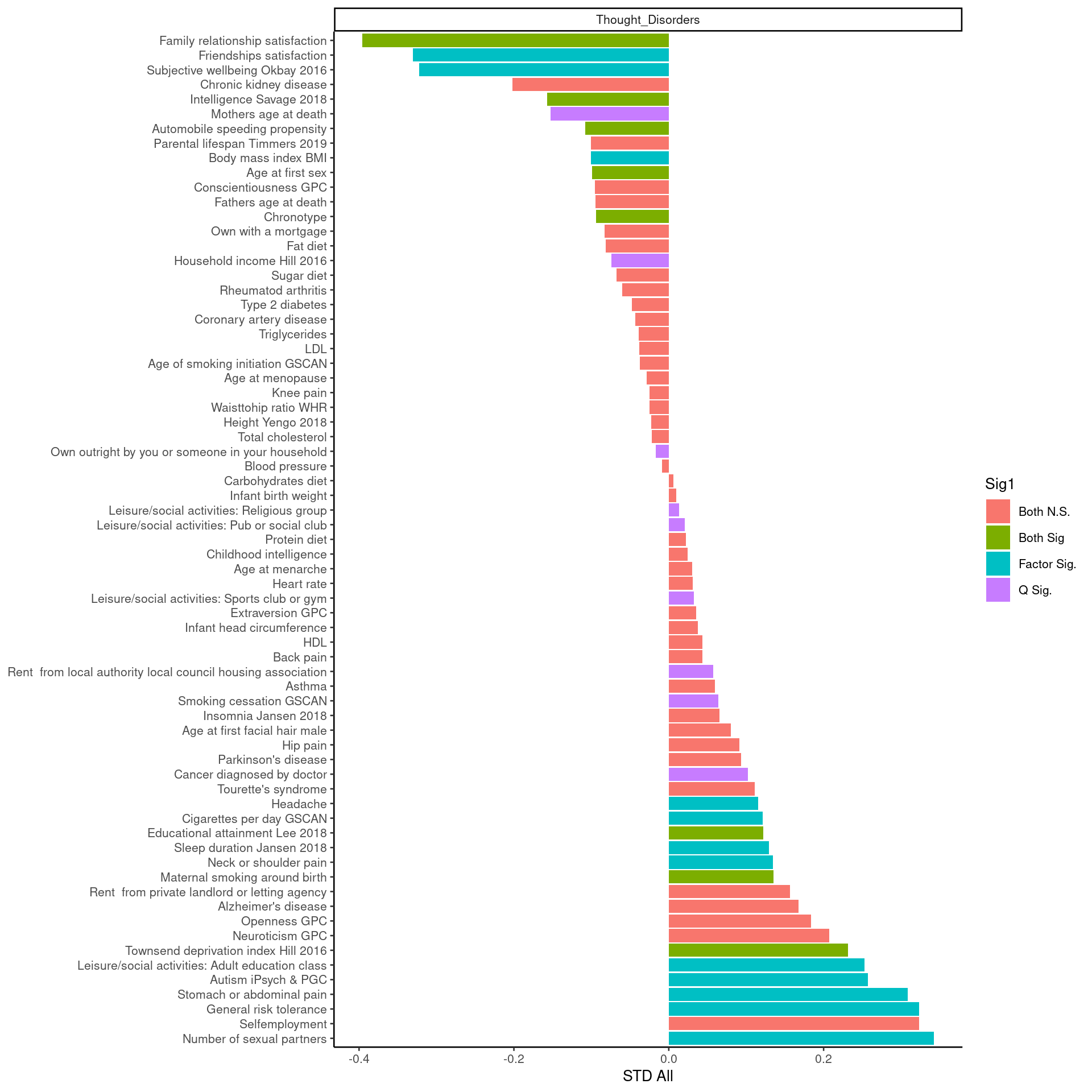

**Supplementary Fig.16. *Q*_Trait_ analyses for the thought disorders factor from the phenotypic correlated factors model.** The significance threshold is set at p < 0.05/69 (the number of external traits). Both N.S. (no factor or *Q* hits). Both Sig. (Significant factor and *Q* hit). *Q* Sig. (Significant *Q* hit, but not factor hit). SNP Sig. (Significant factor hit, but not *Q* hit). A significant *Q* hit indicates that the pattern of genetic correlations between the indicators and the external trait is not well accounted for by the factor.

In the *Q_Trait_* analyses of 68 external traits on the Internalizing factor, 22 out of the 27-factor hits (82%) were also *Q* hits (**Supplementary Fig.17**), suggesting that the pattern of associations between the individual disorders and the external traits is not well accounted for by the Internalizing factor. However, to ensure that the high number of *Q_Trait_* hits reflect factor heterogeneity and did not result from the high-powered nature of the Internalizing factor with its 6 indicators, we created a reduced internalizing factor from indicators that were less genetically correlated (e.g., DEP, PTSD, and OCD) to obtain a less powered Internalizing factor that conserved its heterogeneous nature and still fit the data well (CFI = 0.972, SRMR = 0.089; **Supplementary Fig.18**). Although PTSD and SUI are similarly genetically correlated with DEP, we chose PTSD because it has lower statistical power. Loadings from the full and reduced models were consistent. In the reduced Internalizing model, 16 out of the 24-factor hits (47%) were also *Q* hits and the *Q_Trait_* mean χ^2^ decreased from 46.29 to 14.01, whereas the association mean χ^2^ was consistent across full (17.91) and reduced models (19.26). This suggests that the same amount of signal was captured by the full and reduced Internalizing model. Therefore, the high number of *Q_Trait_* hits in the full model likely stems from the high power of the factor. Given that there were no significant *Q_SNP_* hits (**Table 2**), the internalizing factor appears biologically meaningful as SNP-level (see **Supplementary Fig.19** for Manhattan plot).

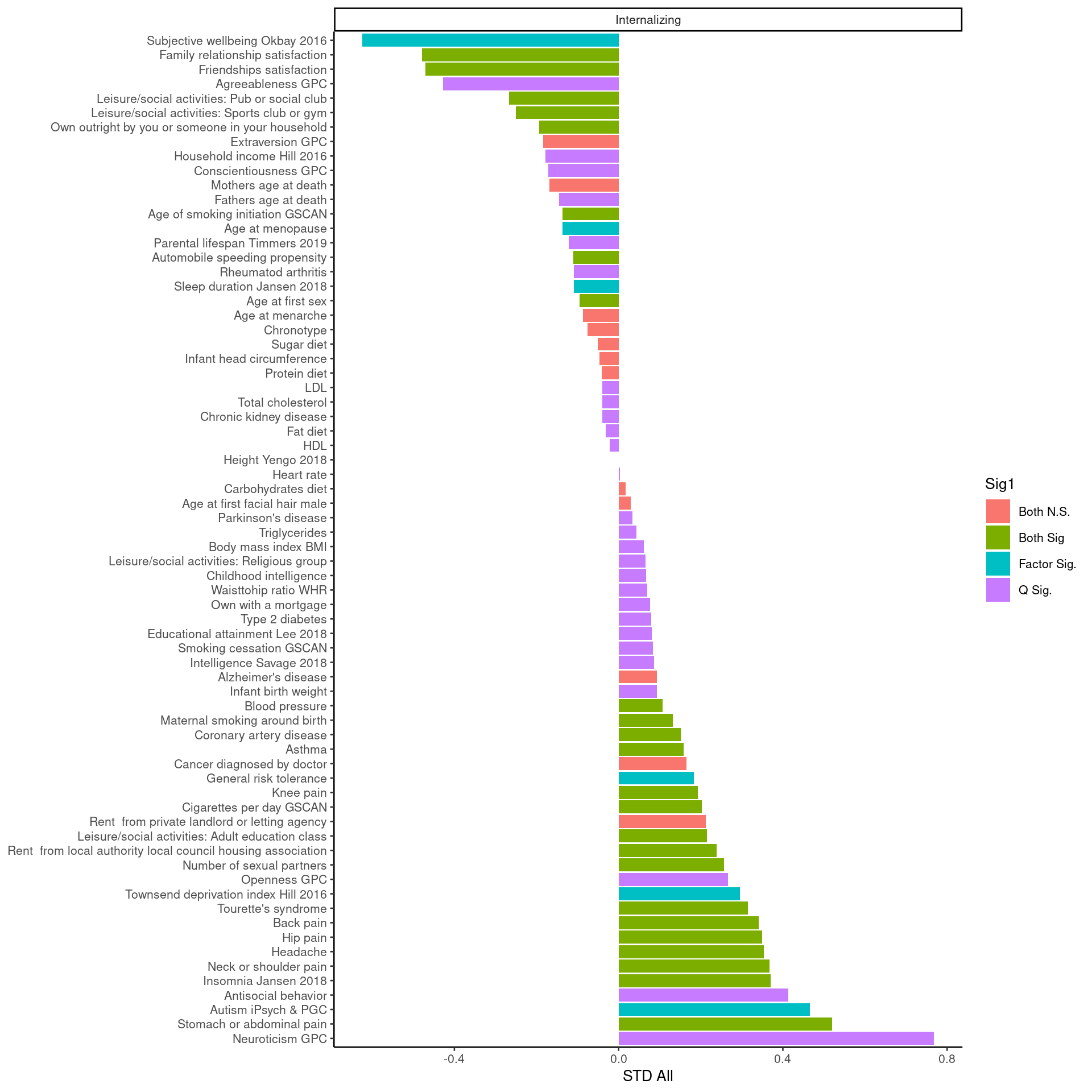

**Supplementary Fig.17. *Q*_Trait_ analyses for the internalizing factor from the phenotypic model.** The significance threshold is set at p < 0.05/70 (the number of external traits). Both N.S. (no factor or Q hits). Both N.S. (no factor or *Q* hits). Both Sig. (Significant factor and *Q* hit). *Q* Sig. (Significant *Q* hit, but not factor hit). SNP Sig. (Significant factor hit, but not *Q* hit). A significant *Q* hit indicates that the pattern of genetic correlations between the indicators and the external trait is not well accounted for by the factor.

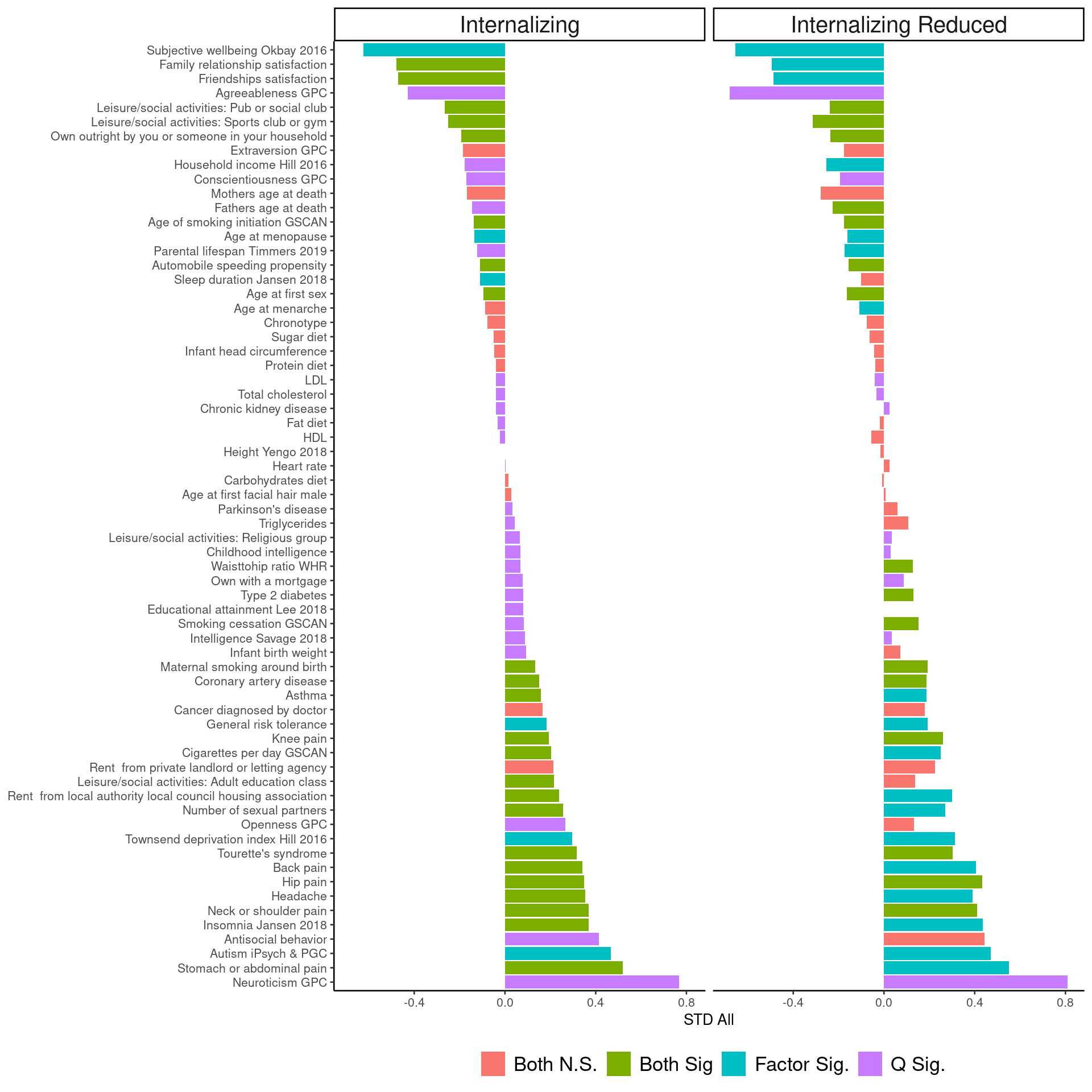

**Supplementary Fig.18. *Q*_Trait_ analyses from the phenotypic model for the internalizing and reduced internalizing factors.** Internalizing factor includes depression, obsessive-compulsive disorder, generalized anxiety, eating disorders, posttraumatic stress disorder, and suicidality, whereas the reduced internalizing factor includes depression, obsessive-compulsive disorder, and posttraumatic stress disorder. The significance threshold is set at p < 0.05/66 (the number of external traits). Both N.S. (no factor or *Q* hits). Both Sig. (Significant factor and *Q* hit). *Q* Sig. (Significant *Q* hit, but not factor hit). SNP Sig. (Significant factor hit, but not *Q* hit). A significant *Q* hit indicates that the pattern of genetic correlations between the indicators and the external trait is not well accounted for by the factor.

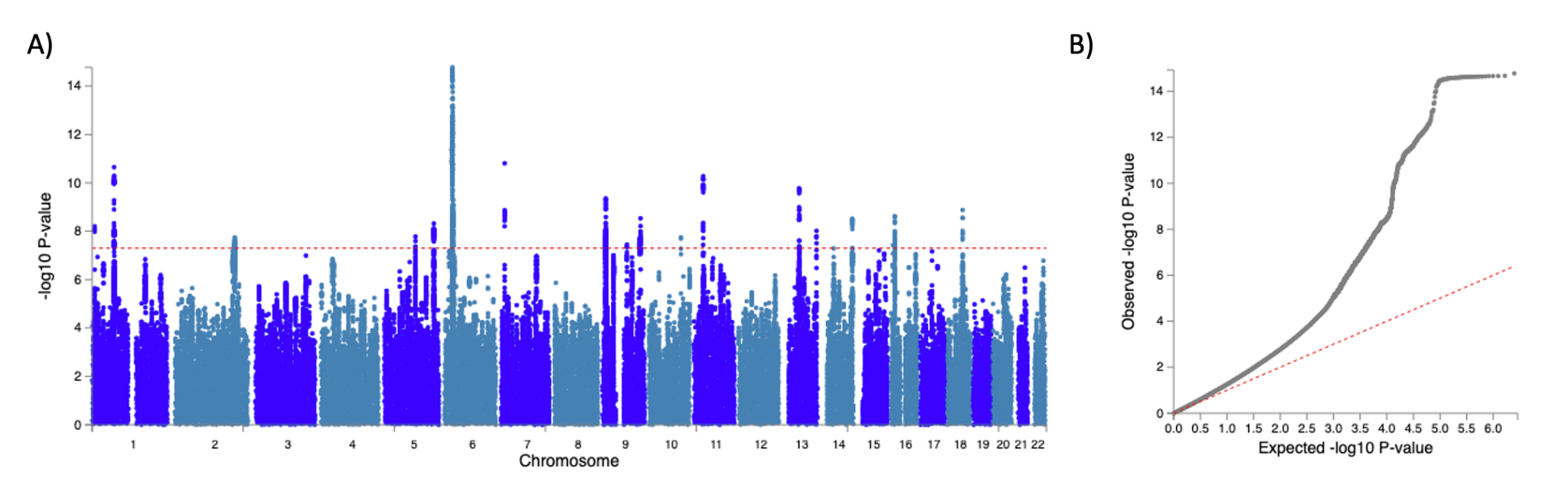

**Supplementary Fig.19. Manhattan plot (A) and QQ-plot (B) of the internalizing disorders factor from the phenotypic correlated factors model.** Manhattan plots: blue "suggestive" line at -log10(1e-5) and red "genome-wide significant" at -log10(5e-8). Miami plot was not provided given that there were no significant *Q_SNP_* hits.

Although there were two factor hits and three *Q* hits in the *Q_Trait_* analyses (**Supplementary Fig.20**), little can be said about the heterogeneity of the externalizing factor because the GWAS had low statistical power and no significant factor or *Q_SNP_* hits (**Table 2**).

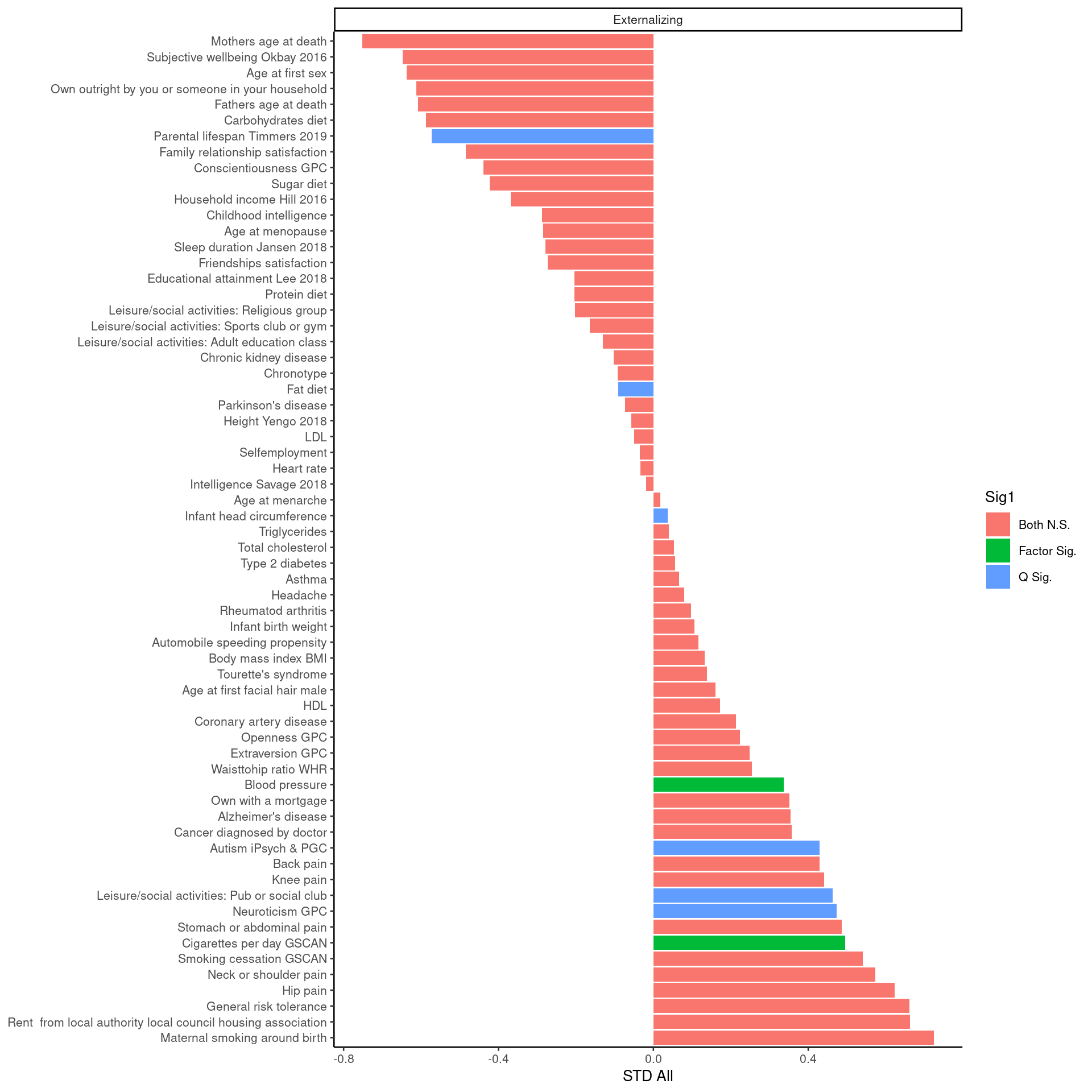

**Supplementary Fig.20. *Q*_Trait_ analyses for the externalizing factor from the phenotypic model.** The significance threshold is set at p < 0.05/70 (the number of external traits). Both N.S. (no factor or *Q* hits). Both Sig. (Significant factor and *Q* hit). *Q* Sig. (Significant *Q* hit, but not factor hit). SNP Sig. (Significant factor hit, but not *Q* hit). A significant *Q* hit indicates that the pattern of genetic correlations between the indicators and the external trait is not well accounted for by the factor.

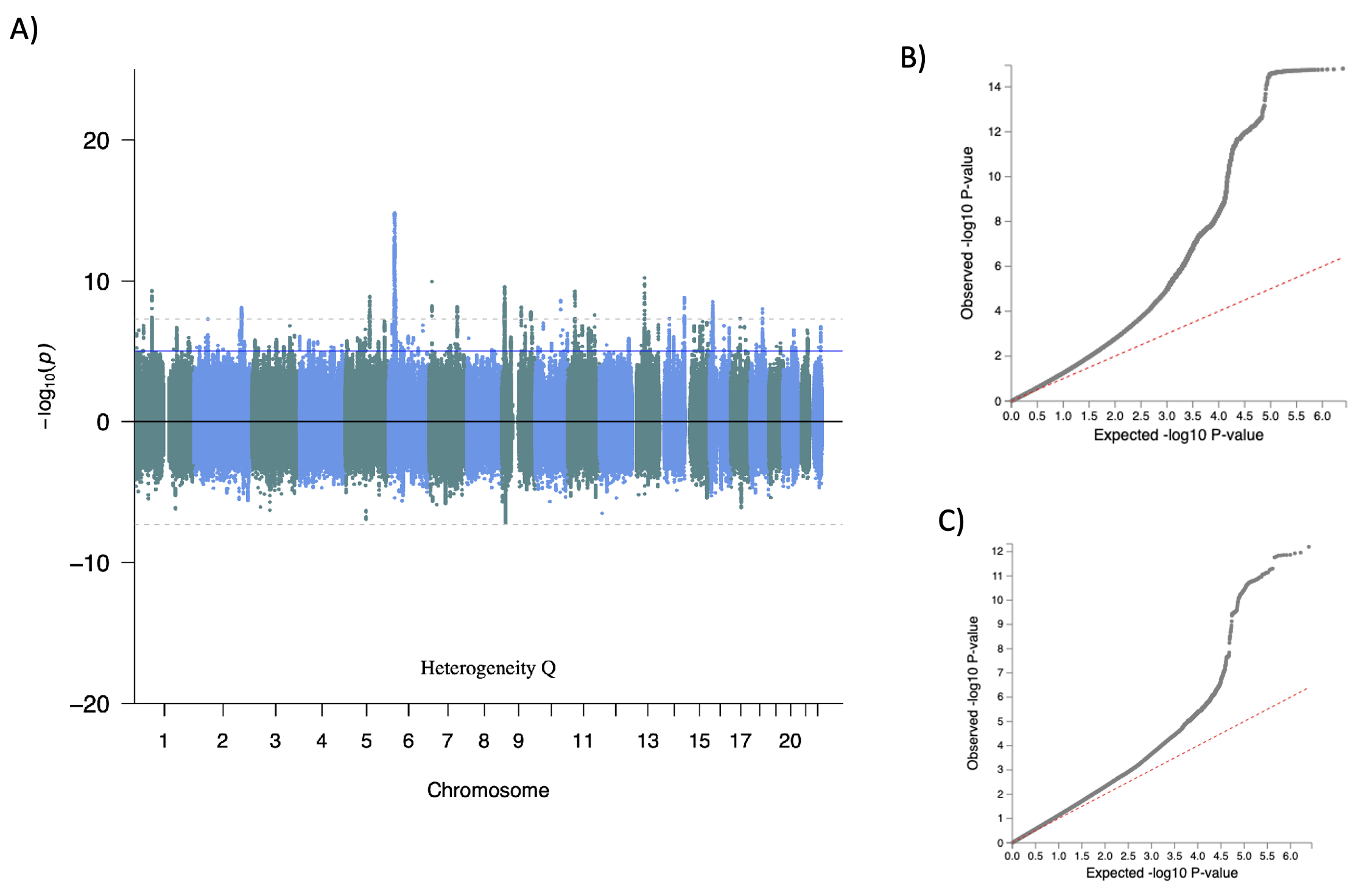
**Supplementary Fig.21. Miami plot (A), factor QQ-plot (B), and *Q*_SNP_** **QQ-plot (C) of the internalizing factor from the genetically informed correlated factors model.** Dotted grey "genome-wide significant" line at -log10(5e-8) and blue "suggestive" line at -log10(1e-5). Panel (A) top: first-order factor Genome-wide Association Study (GWAS) results. Panel (A) bottom: heterogeneity *Q* GWAS.

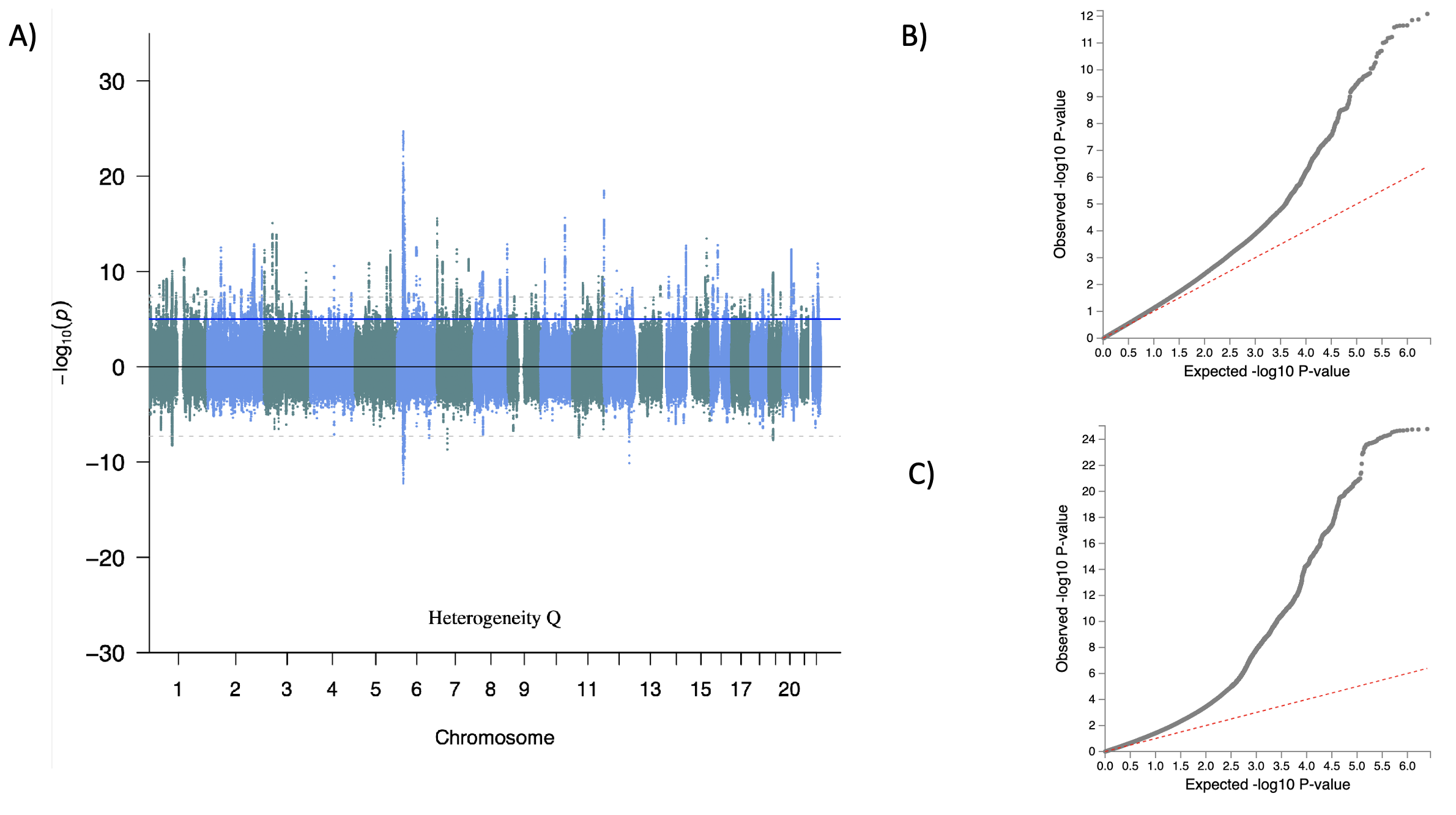

**Supplementary Fig.22. Miami plot (A), factor QQ-plot (B), and *Q*_SNP_** **QQ-plot (C) of the thought disorders factor from the genotypic correlated factors model.** Dotted grey "genome-wide significant" line at -log10(5e-8) and blue "suggestive" line at -log10(1e-5). Panel (A) top: first-order factor Genome-wide Association Study (GWAS) results. Panel (A) bottom: heterogeneity *Q* GWAS.

#### 7. **Neural Correlates of Psychopathology**

To evaluate associations between the PGIs of psychopathology and brain measures, we examined the association between cortical surface areas and cortical mean thicknesses with each polygenic index (**Supplementary Fig. 23-24**). The brain-polygenic index association was highly similar between the first- and second-order polygenic indices (r = 0.982 for surface areas, r = 0.961 for mean thicknesses). In light of the high correlation between the first- and second-order *p* factor polygenic indices, we report the association of the INT polygenic index, the SU polygenic index, and the TD polygenic index with the first-order *p* factor polygenic index.

In mean thicknesses, the first-order *p* factor and INT brain-polygenic index associations are highly positively correlated (r = =0.751), whereas the first-order *p* factor and EXT brain-polygenic index were moderately positively correlated (r = =0.298) and first-order *p* factor and TD brain-polygenic index associations were not significantly correlated (r = =0.128). In surface areas, the first-order *p* factor and INT brain-polygenic index associations are highly positively correlated (r = =0.759), whereas the first-order *p* factor and EXT brain-polygenic index and the first-order *p* factor and TD brain-polygenic index associations were moderately positively correlated (r = =0.492 and r = =0.271, respectively).

The INT_p_ polygenic index positively predicted mean cortical thicknesses that were negatively associated with the TD_p_ polygenic index (r = -0.395), suggesting that the genes associated with Internalizing have the opposite effect to those associated with thought disorders. The polygenic index-brain associations of INT_p_ with TD_p_ and of INT_p_ with EXT_p_ were not correlated. As for surface areas, the polygenic index-brain associations across the INT_p_, TD_p_, and EXT_p_ factors were not significantly correlated. Our findings support distinct associations between psychiatric liabilities and cerebral measures.

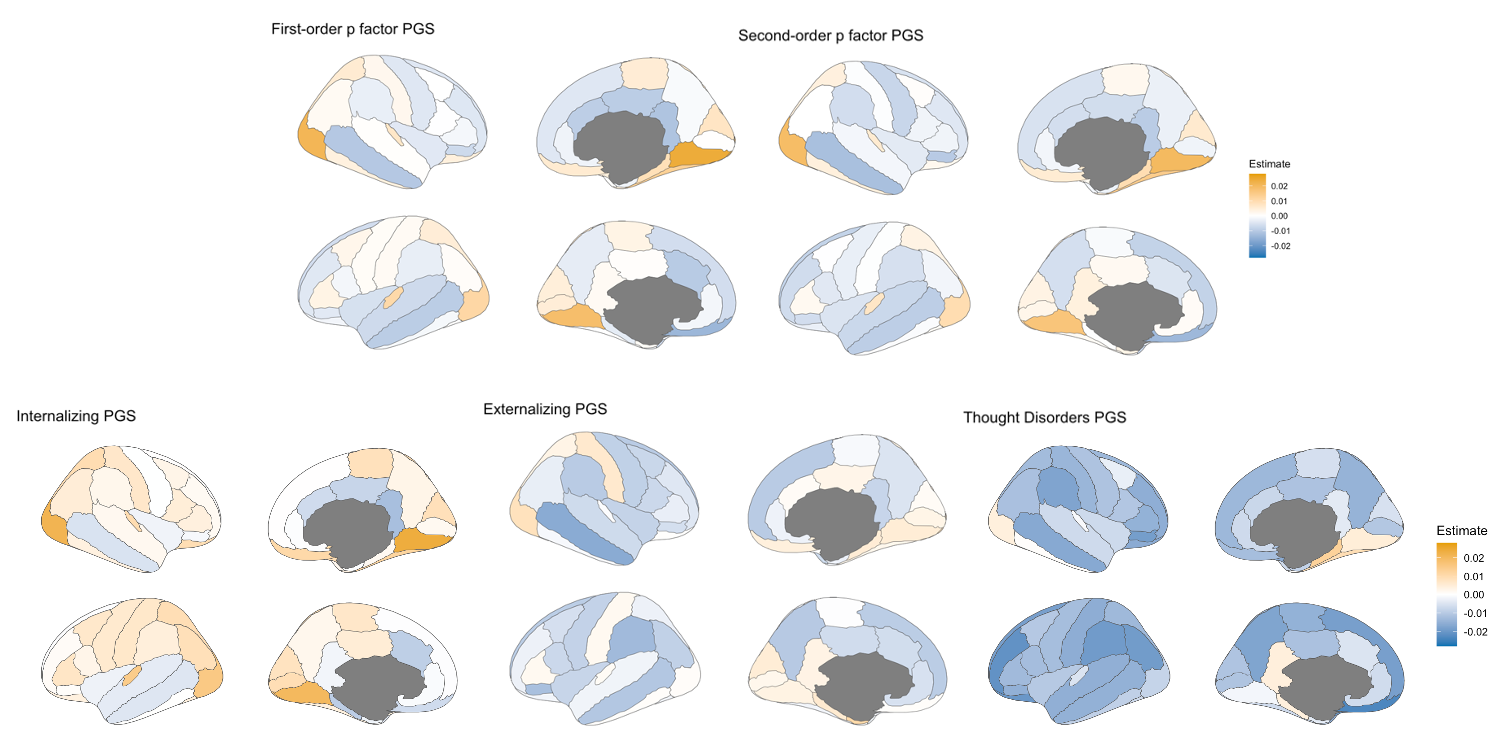

**Supplementary Fig.23. Standardized estimate of the polygenic index (PGI) effect across cortical mean thicknesses**. Blue regions indicate that a greater polygenic index is associated with less cortical surface area in that region, whereas orange regions indicate that a greater polygenic index is associated with greater cortical surface area in that region. All regions regardless of significance. Not adjusted for global brain size. See **Supplementary Table 13** for regression results and significance testing. polygenic indices were residualized for year of birth and the first 40 principal components of the genetic data. Models with cerebral measures included sex, age (at the scanner visit), age^2^, + sex*age, sex*age^2^, and scanner site as covariates. First-order *p* factor: *p* factor from the first-order *p* factor model. Second-order *p* factor: *p* factor from the second-order *p* factor model.

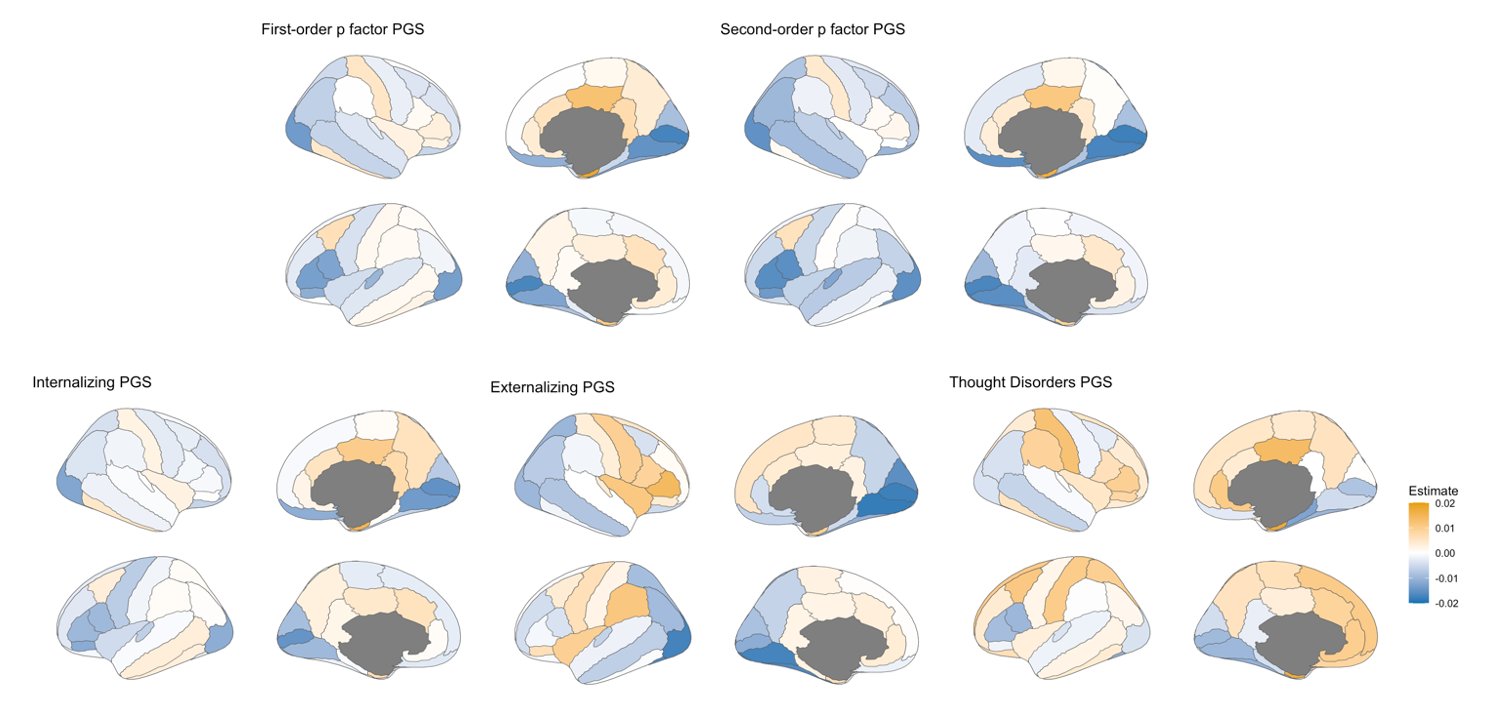

**Supplementary Fig.24. Standardized estimate of the polygenic index (PGI) effect across cortical surface areas**. Blue regions indicate that a greater polygenic index is associated with less cortical surface area in that region, whereas orange regions indicate that a greater polygenic index34 is associated with greater cortical surface area in that region. All regions regardless of significance. Not adjusted for global brain size. See **Supplementary Table 13** for regression results and significance testing. polygenic indices were residualized for year of birth and the first 40 principal components of the genetic data. Models with cerebral measures included sex, age (at the scanner visit), age^2^, + sex*age, sex*age^2^, and scanner site as covariates. First-order *p* factor: *p* factor from the first-order *p* factor model. Second-order *p* factor: *p* factor from the second-order *p* factor model.

8. **Phenome-wide association study (PheWAS) Results**

To evaluate associations between the PGIs of psychopathology and a wide variety of medical outcomes, we performed a PheWAS for each PGI in genotyped individuals of European ancestry in the Mass General Brigham Biobank (MGBB^4^) biorepository with a logistic regression fit to 1,819 case/control disease phenotype.

The effects of the first-and second-order *p* factor PGIs on the case/control disease phenotypes were correlated at 0.967. The effects of the TD_p_ and TD_g_ PGI on the case/control disease phenotypes were correlated at 1 and those of the INT_p_ and INT_g_ PGI were correlated at 0.985. Generally, the *p* factor PGIs were more broadly associated with medical outcomes. For instance, the second-order *p* PGI was associated with 626 (36%) medical outcomes, whereas the INT_p_ PGI was associated with 512 (29%) medical outcomes (absolute mean effect sizes of 0.134 and 0.144, respectively; **Supplementary Tables 14)**.

Of the 5% of medical outcomes that were only significantly associated with the *p* factor PGI, the absolute mean difference in estimate between the *p* factor and the factor with the highest estimate was 0.027, suggesting that the effect size of these associations may become significantly associated with the domain-level transdiagnostic factors PGIs if the analyses were better powered. In contrast, the effect sizes of the medical outcomes that were associated with the domain-level transdiagnostic factors, but not the *p* factor PGI, were generally larger: the absolute mean difference in effect size between the *p* factor and the TD factor PGI was 0.087, it was 0.055 for INT, 0.068 for COMP and 0.089 for EXT.

The correlation between PGIs was more variable across PGIs from the genotypic and phenotypic models (**Supplementary Fig.25-27**). For instance, the effects of the INT_p_ and TD_p_ PGI were generally correlated at 0.403. This was largely driven by the significant negative association (r = -0.253) between their effects on injuries and poisonings (**Supplementary Fig.25a**). Therefore, the effects of the PGIs across disease phenotypes vary across PGIs and disease phenotypes.

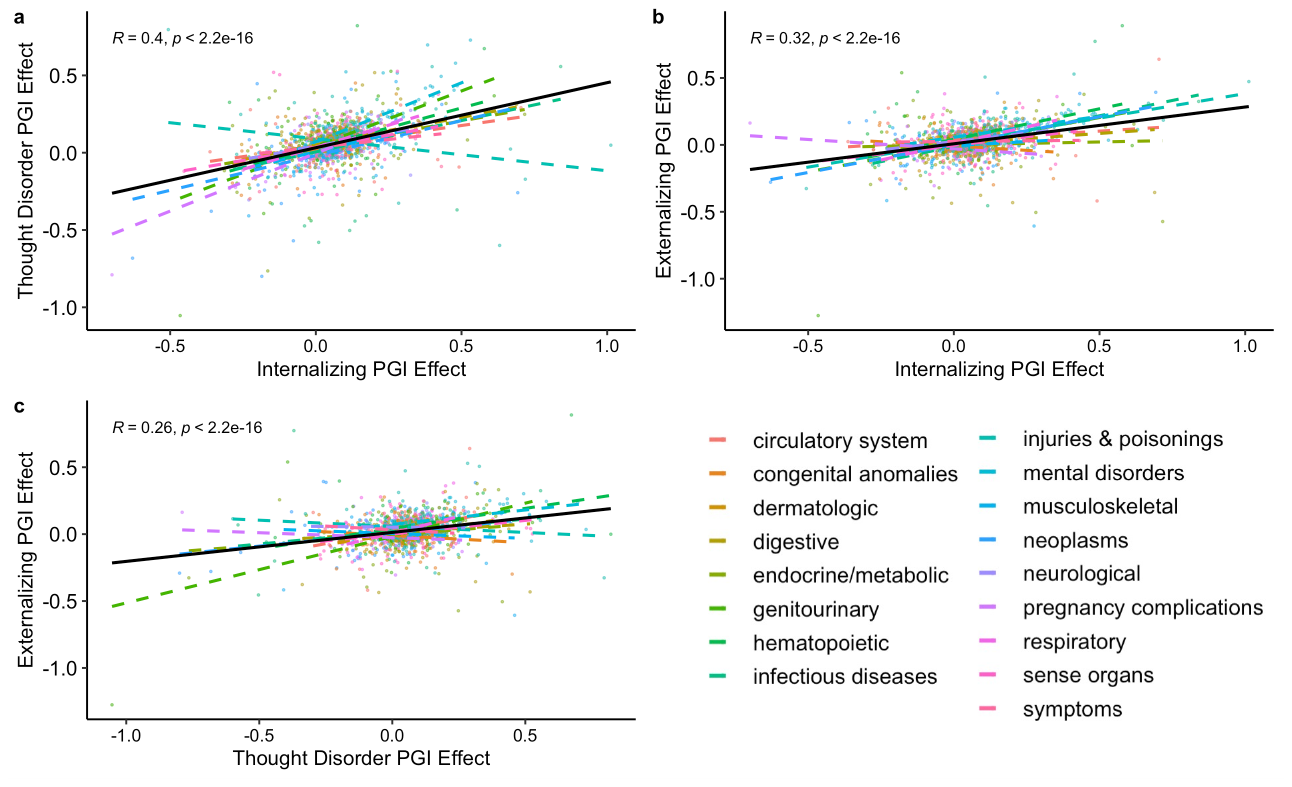

**Supplementary Fig.25. Correlation between the effects of the polygenic index (PGI) on the 1, 817 disease phenotypes across psychopathology factors of the phenotypic correlated factors model*.*** Solid black line reflects the average correlation across all types of medical outcomes.

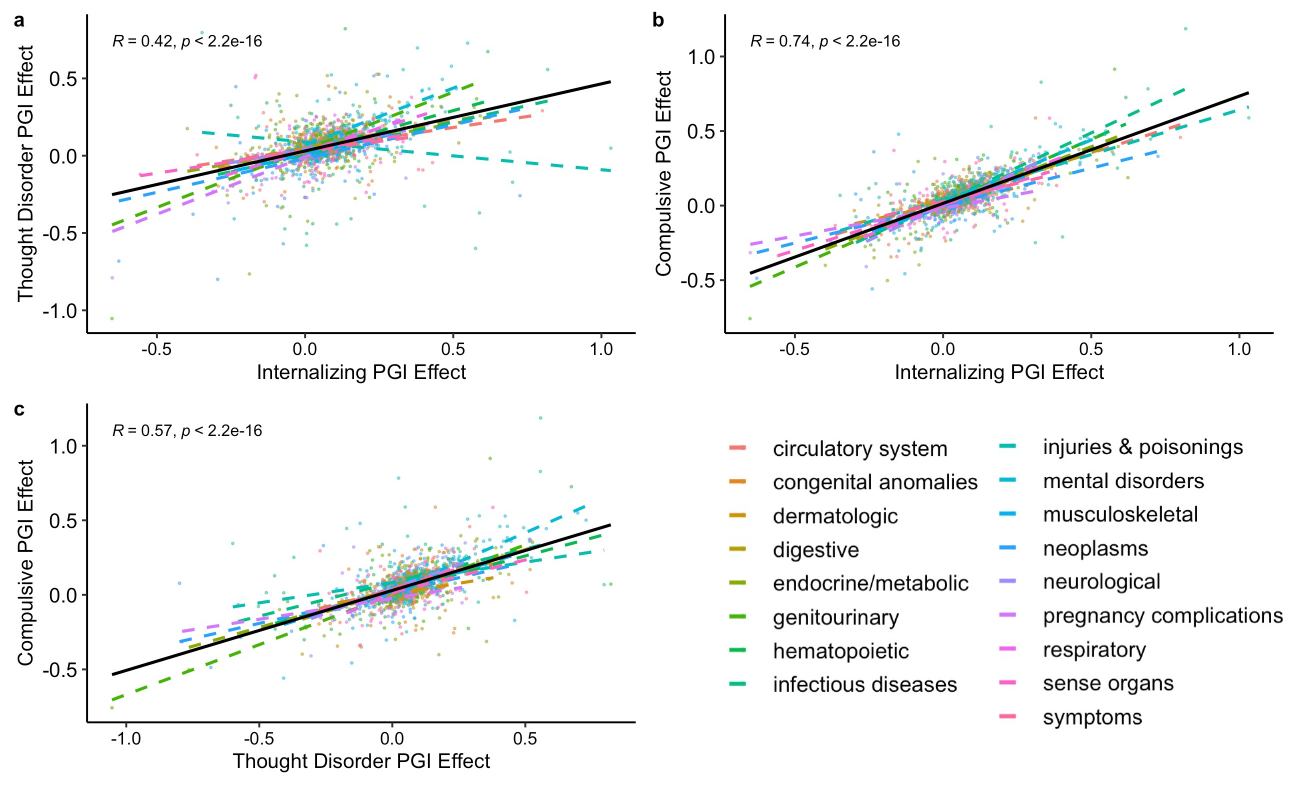

**Supplementary Fig.26. Correlation between the effects of the polygenic index (PGI) on the 1, 817 disease phenotypes across psychopathology factors from the genotypic correlated factors Model.** Solid black line reflects the average correlation across all types of medical outcomes.

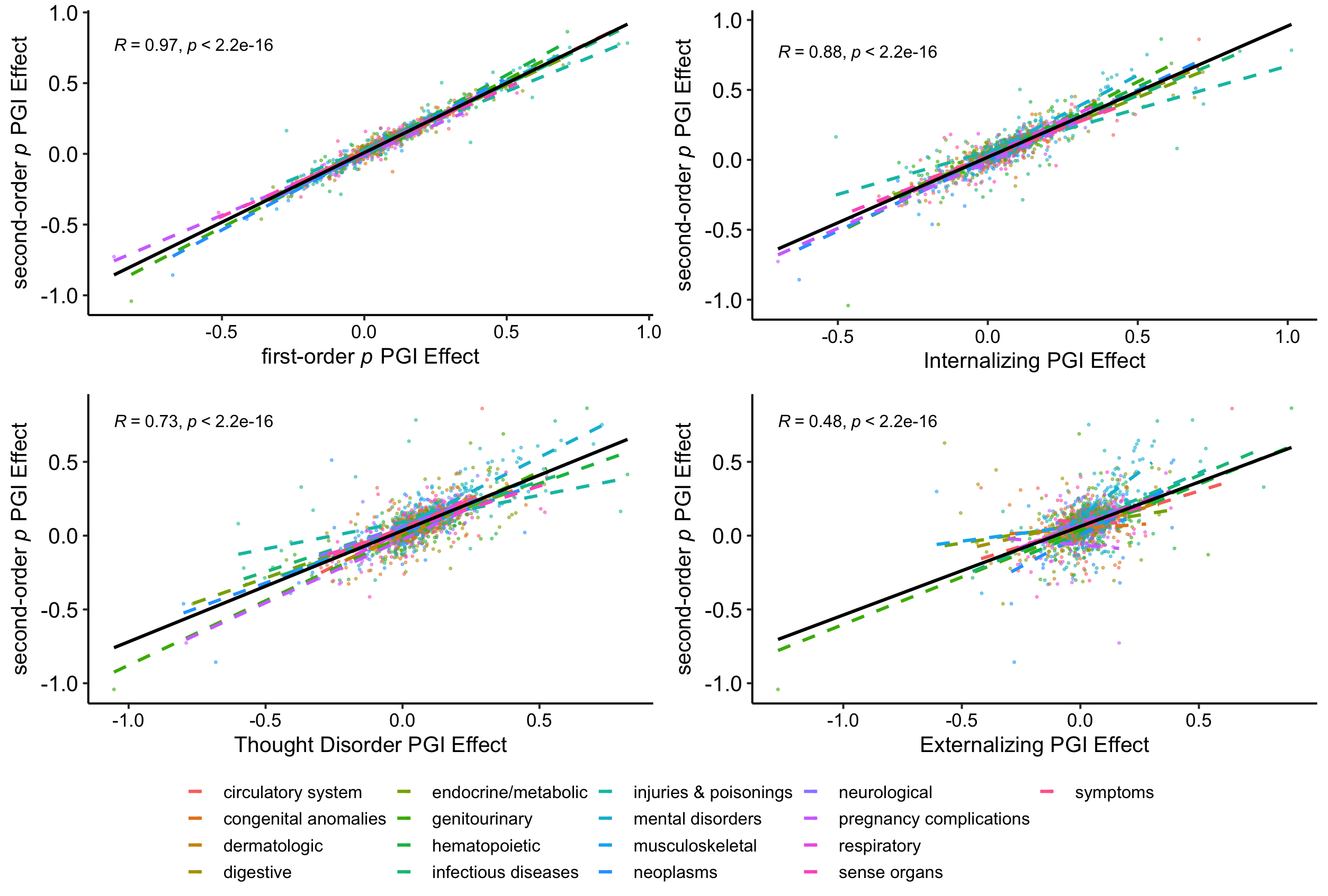

**Supplementary Fig.26. Correlation between the Ssecond-order *p* factor Polygenic Index (PGI) effects on the 1,817 disease phenotypes and the PGI effects from the phenotypic correlated factors model.** Solid black line reflects average correlation across all types of medical outcomes.
